# Microbial context predicts SARS-CoV-2 prevalence in patients and the hospital built environment

**DOI:** 10.1101/2020.11.19.20234229

**Authors:** Clarisse Marotz, Pedro Belda-Ferre, Farhana Ali, Promi Das, Shi Huang, Kalen Cantrell, Lingjing Jiang, Cameron Martino, Rachel E. Diner, Gibraan Rahman, Daniel McDonald, George Armstrong, Sho Kodera, Sonya Donato, Gertrude Ecklu-Mensah, Neil Gottel, Mariana C. Salas Garcia, Leslie Y. Chiang, Rodolfo A. Salido, Justin P. Shaffer, MacKenzie Bryant, Karenina Sanders, Greg Humphrey, Gail Ackermann, Niina Haiminen, Kristen L. Beck, Ho-Cheol Kim, Anna Paola Carrieri, Laxmi Parida, Yoshiki Vázquez-Baeza, Francesca J. Torriani, Rob Knight, Jack A. Gilbert, Daniel A. Sweeney, Sarah M. Allard

## Abstract

Synergistic effects of bacteria on viral stability and transmission are widely documented but remain unclear in the context of SARS-CoV-2. We collected 972 samples from hospitalized patients with coronavirus disease 2019 (COVID-19), their health care providers, and hospital surfaces before, during, and after admission. We screened for SARS-CoV-2 using RT-qPCR, characterized microbial communities using 16S rRNA gene amplicon sequencing, and contextualized the massive microbial diversity in this dataset through meta-analysis of over 20,000 samples. Sixteen percent of surfaces from COVID-19 patient rooms were positive, with the highest prevalence in floor samples next to patient beds (39%) and directly outside their rooms (29%). Although bed rail samples increasingly resembled the patient microbiome over time, SARS-CoV-2 was detected less there (11%). Despite viral surface contamination in almost all patient rooms, no health care workers contracted the disease, suggesting that personal protective equipment was effective in preventing transmissions. SARS-CoV-2 positive samples had higher bacterial phylogenetic diversity across human and surface samples, and higher biomass in floor samples. 16S microbial community profiles allowed for high SARS-CoV-2 classifier accuracy in not only nares, but also forehead, stool, and floor samples. Across distinct microbial profiles, a single amplicon sequence variant from the genus *Rothia* was highly predictive of SARS-CoV-2 across sample types and had higher prevalence in positive surface and human samples, even compared to samples from patients in another intensive care unit prior to the COVID-19 pandemic. These results suggest that bacterial communities may contribute to viral prevalence both in the host and hospital environment.

**One Sentence Summary:** Microbial classifier highlights specific taxa predictive of SARS-CoV-2 prevalence across diverse microbial niches in a COVID-19 hospital unit.

## Introduction

Severe acute respiratory syndrome coronavirus 2 (SARS-CoV-2) is the causative agent of a novel infectious disease, COVID-19, that has reached pandemic proportions. COVID-19 was first detected in Wuhan, China, in patients with pneumonia in December 2019. This pandemic has been characterized by sustained human to human transmission and it has caused more than 44 million cases and over 1.2 million deaths worldwide (as of 1 November 2020, WHO report). The United States now has the largest number of cases worldwide at over 11 million as of November 20th, 2020 *(1)*. COVID-19 is primarily transmitted via either respiratory droplets or aerosols produced by an infected person and inhaled by another individual. Other routes of transmission have also been proposed including fecal oral transmission *(2, 3)* and fomite transmission *(4)* although the relative importance of various transmission routes is uncertain *(5–8)*. The potential role of fomite transmission is especially concerning as SARS-CoV-2 has been detected on a variety of surfaces including plastic, stainless steel, cardboard, and copper, and in aerosols *(9)*. A more comprehensive understanding of what influences SARS-CoV-2 stability, transmission, and infectivity is crucial to implementing effective public health measures.

Viruses exist in a complex microbial environment, and virus-bacterial interaction has been increasingly documented in humans. In the animal microbiome, the gastrointestinal tract contains the highest amount of bacteria and many virus-bacterium interaction studies have therefore focused on enteric viruses. Gut bacteria have been shown to directly modulate enteric virus infectivity via improving thermostability *(10)*, increasing environmental stability *(11)*, and encouraging viral genetic diversity and fitness *(12)*. Virus-bacterium interactions have also been observed in upper-respiratory tract infections including influenza A *(13, 14)* and oral human papillomavirus infection *(15)*. Most recently, prevalent bacteria in the human microbiome have been demonstrated to alter the human glycocalyx thereby modulating the ability of SARS-CoV-2 to bind host cells *(16)*. Given the nature of known virus-bacterium interactions, we hypothesized that virus-bacterium interactions may also exist in indoor spaces (the ‘built environment’).

The risk of contracting SARS-CoV-2 is higher indoors than outdoors particularly in poorly ventilated areas *(17)*, and the built environment has a distinct microbiome *(18)*. The built environment microbiome is usually dominated by human-associated microbes *(19)*, and it is estimated that humans shed approximately 37 million bacterial genomes per hour into their built environments *(20)*. In a study following the building of a new hospital, we discovered that the indoor spaces were colonized with microbes from patients and health care workers, and metagenomic analysis was used to infer transmission between occupants via surface transmission *(21)*. To test whether specific bacterial taxa in the host or built environment influence SARS-CoV-2 persistence, we collected samples from hospital surfaces, patients, and health care workers in the intensive care unit (ICU) and medical-surgical floor during the onset of the COVID-19 outbreak and screened for viral presence and microbial context.

## Results

### SARS-CoV-2 detection across surfaces and patient samples

Sample collection for SARS-CoV-2 detection is typically performed using viral transport media containing fetal bovine serum and a cocktail of antibiotics, which could negatively influence studies of bacteria and other microbes *(22, 23)*. For this study, swab samples were stored in 95% EtOH, in order to inactivate the virus for safe transportation *(24)* while stabilizing the microbial community *(25)*. A total of 972 samples were collected longitudinally from 16 patients with clinical laboratory confirmed SARS-CoV-2 infection (118 samples), 10 health care workers assigned to these patients (113 samples), and 734 hospital surfaces either inside or immediately outside of the patients’ rooms over the span of two months (Fig. 1A).

**Figure 1.**
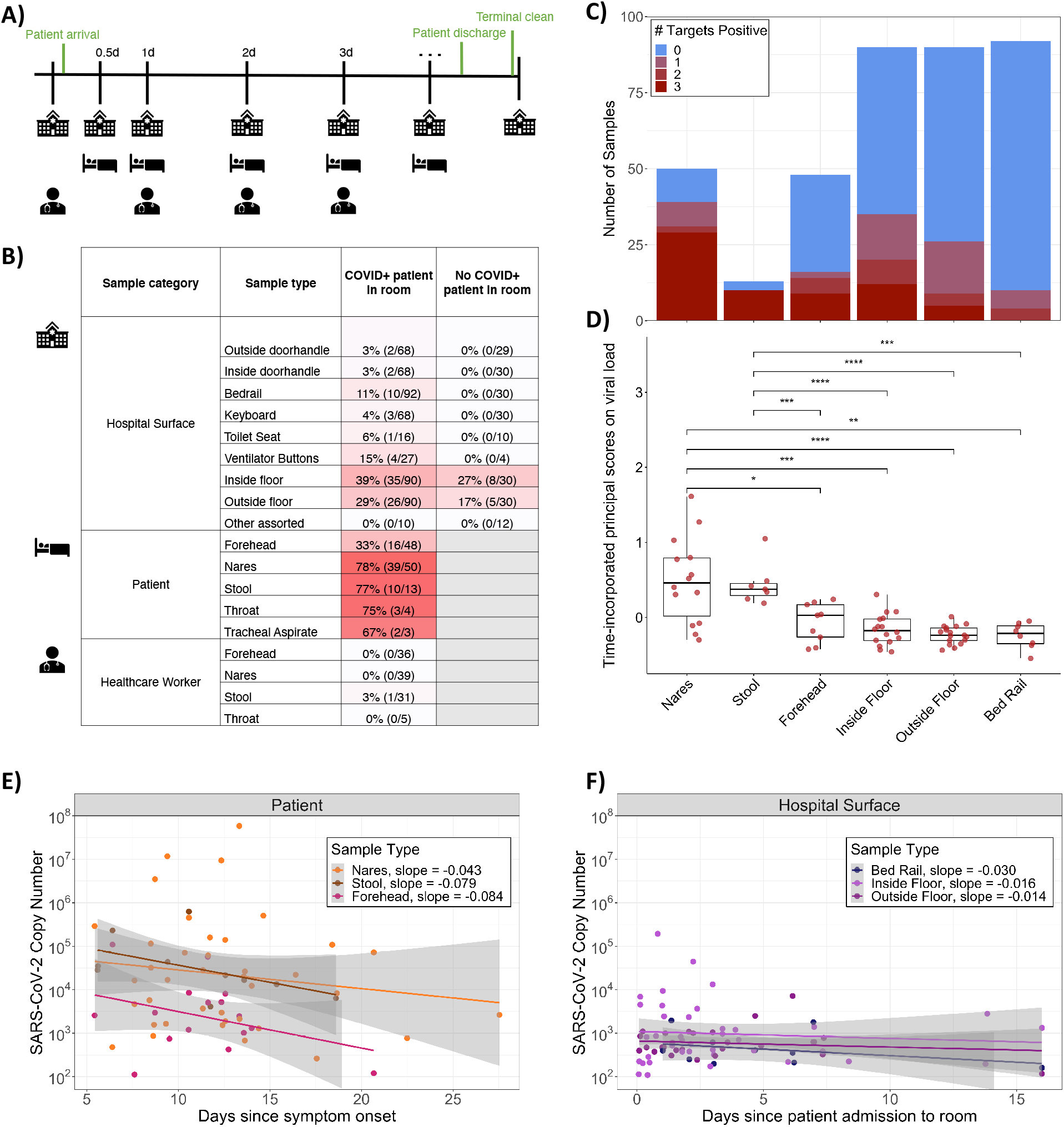
Summary of SARS-CoV-2 detection in the dataset. **A**) Schematic diagram of the experimental design highlighting the time frame for sample collection across sample types. **B**) Percent and number of COVID-positives for each sample type collected from rooms occupied or not occupied by COVID-19 patients. Not occupied includes both post-cleaning rooms and rooms currently occupied by a patient negative for COVID-19. **C)** Number of samples and SARS-CoV-2 screening results for 3 gene targets (N1, N2, and E-gene). **D**) Boxplot of time-incorporated principal scores on viral load for different sample types. Each dot represents the functional principal component score for each viral load trajectory over time, which was estimated from sparse functional principal components analysis on viral load over time; *p<0.05, **p<0.01, ***p<0.001, ****p<0.0001, Wilcoxon signed-rank test. **E**) Viral load per swab relative to date of symptom onset across COVID-19 patient sample types, where only sample types with both n positive>10 and % positive>10% are included. (**F**) Viral load per swab relative to date of room admission across hospital surface sample types, where samples from rooms occupied by a COVID-19 patient at the time of sampling are included. Again, sample types with both n>10 and % positive>10% are included.

The 16 patients enrolled in this study ranged from age 20 to 84, with a median age of 49.5 (Fig S1). 31% were female and 69% were male, consistent with reports that men tend to experience more severe COVID-19 symptoms *(26)*. Of the patients for whom antibiotic treatment information was collected, 77% were on antibiotics, of which 80% were taking more than one antibiotic. The number of days spent in the hospital ranged from 1 to 25, with a median stay of 9 days.

Each sample was screened for the presence of SARS-CoV-2 using three distinct primer/probe sets: the U.S. Center for Disease Control N1 and N2 targets, and the World Health Organization E-gene target (see methods). The US Food and Drug Administration has issued Emergency Authorization for more than 150 RT-qPCR assays for the detection of SARS-CoV-2, the majority of which define a positive result as amplification in a single target *(27)*. Accordingly, we designated samples as positive if at least one out of three targets amplified with a Ct value below 40. Serial dilutions of quantified virus amplicons were included in each RT-qPCR plate in order to extrapolate the viral load of each sample. Of the surfaces sampled, 13.1% were positive for SARS-CoV-2, including those touched primarily by health care workers (keyboard, ventilator buttons, door handles inside, and outside the rooms) and those directly in contact with the patient (toilet seats, bed rails). Of the patients enrolled in the study, we collected at least one positive sample from 15/16 patients (nares, forehead, or stool) and from 14/15 associated hospital rooms. In rooms where patient samples were not available, surfaces screened positive at least once for 6/6 COVID-19 rooms and 4/5 non-COVID-19 rooms.

Floor samples had the highest positivity rates (36% of samples collected from the floor near the patients’ bed, i.e. “Inside Floor”, and 26% of samples collected from the floor immediately outside of the patient room, i.e. “Outside Floor”) (Fig. 1B, Fig. S2). In some cases, SARS-CoV-2 was detected on the floors of rooms with patients who tested negative for COVID-19 and in rooms that had been cleaned following COVID-19 patient occupancy (Fig. 1B, Fig. S3B). Most of the positive surface samples amplified only one or two out of the three SARS-CoV-2 targets (Fig. 1C) and had significantly lower viral load over time compared to patient nares and stool samples (*p*<0.003, non-parametric test from sparse functional principal components analysis) *(28)*, but similar viral load to patient forehead samples (Fig. 1D).

SARS-CoV-2 viral load tended to decrease in patients over time (Fig. 1E) but was detectable in patient nares up to 27 days after symptom onset. Trajectories of viral load varied for different patients (Fig. S3). For a COVID-19-positive patient’s stay, viral load also tended to decrease slightly on hospital surfaces including bed rails and floor samples but remained detectable up to 16 days after patient admission (Fig. 1F).

Of 113 health care worker samples, only one stool sample amplified for one of the three viral targets. No other samples collected from this health care worker, and no samples from any other health care worker treating COVID-19 patients had any viral target amplification. Moreover, all health care workers in this study did not have detectable serum antibodies against SARS-CoV-2.

### Diverse microbial context of SARS-CoV-2

16S V4 rRNA gene amplicon (16S) sequencing was performed and a total of 589 out of the 972 samples passed quality filtering (see methods). Most of the sample dropouts were low biomass samples from surfaces in the built environment (49% of hospital surface samples compared to 9% of human samples). Fewer samples that failed 16S sequencing were SARS-CoV-2 positive (6.7%) compared to samples that sequenced successfully (23.9%). A meta-analysis with samples from the Earth Microbiome Project *(29)*, an intensive care unit microbiome project *(30)*, and a hospital surface microbiome study performed at another hospital *(21)* (a total of 19,947 samples) contextualized the microbial composition of samples from this hospital study and the broad range of microbial diversity covered in this dataset (Fig. 2A). Through source-tracking *(31)* on the meta-analysis we found that floor samples, which cluster separately from the rest of this dataset (Fig. 2C), are similar to built environment samples from previous studies (Fig. S4).

**Figure 2.**
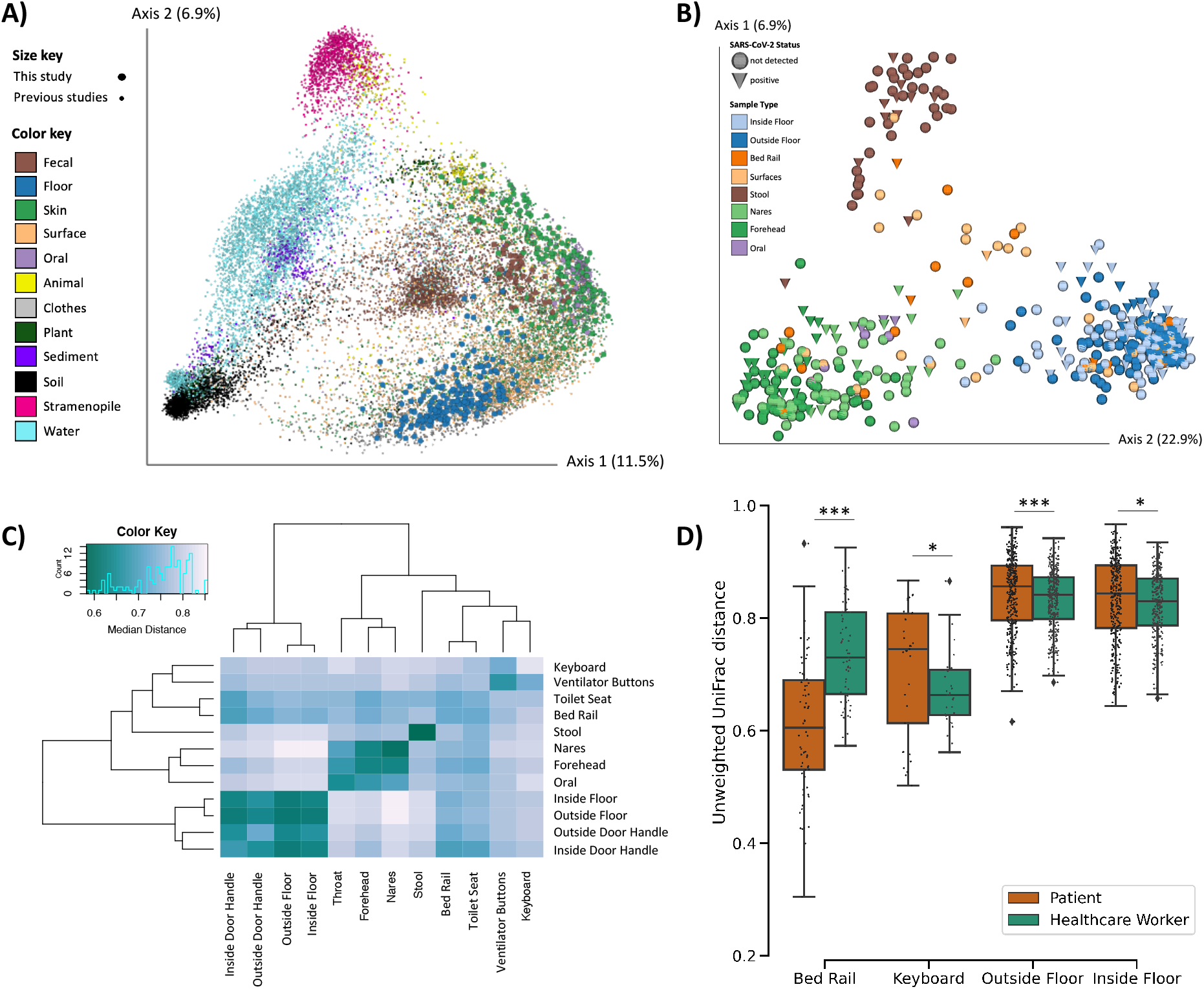
Microbial diversity of SARS-CoV-2 patients, health care workers, and the built environment in COVID-19 units. **A)** Principal Coordinates Analysis (PCoA) of unweighted UniFrac distances comparing the Earth Microbiome Project meta-analysis (n=19,497, small dots) and this study (n=591, large dots). **B)** PCoA of unweighted UniFrac distances in this study. **C)** Heatmap of unweighted UniFrac distance among surface and patient sample types. Diagonal lines represent median distances within individual sample types. **D)** Pairwise unweighted UniFrac distance between the human surface (i.e. forehead and nares) and their paired surface samples. Statistics represent bootstrapped Kruskal-Wallis; *p<0.05, **p<0.01, ***p<0.001.

Beta-diversity estimated using unweighted UniFrac distances *(32)* in this study showed that floor samples, stool samples, and nares/forehead samples formed three distinct clusters with other surfaces falling between the human skin and floor samples (Fig. 2B-C). SARS-CoV-2 viral load was weakly correlated with unweighted UniFrac beta-diversity (PERMANOVA R^2^ <0.01, p-value = 0.043, Fig. S5).

We compared beta-diversity between human samples and paired built environment samples from the patients’ respective hospital rooms. Microbial composition of high touch surfaces routinely used by healthcare workers, such as keyboards and floor samples, were significantly more similar to health care worker samples, whereas samples from bed rails that are not frequently touched by health care workers were significantly more similar to the patient samples (Fig. 2D). Notably, the percent of SARS-CoV-2 positive bed rail samples was lower than floor (11% vs 39%) despite the high similarity of bed rail microbiomes to the corresponding patient microbiomes.

### Longitudinal beta-diversity analysis reveals patient-surface microbial convergence

To account for the longitudinal nature of this dataset, we applied a compositional tensor factorization method implemented through the Gemelli QIIME2 plugin *(33, 34)* (Fig. 3A). *Actinomycetales* and *Bacteroidales* were the most highly ranked taxa driving the separation of patient’s forehead and nares samples from surface samples, separating those two groups along the first principal component axis (PC1). *Bacillales* was also ranked among the top contributors to microbial separation in our dataset and has been successfully used for biocontrol on hospital surfaces *(35–38)*. The log-ratio of *Bacillales* versus *Actinomycetales* and *Bacteroidales* was higher in surface samples compared to human samples (Fig. 3B). The trajectory of this log-ratio showed that with longer hospitalizations, bed rail samples became more similar to patients’ nares and forehead samples. Upon patient discharge and room cleaning, this log-ratio converged back towards floor samples.

**Figure 3.**
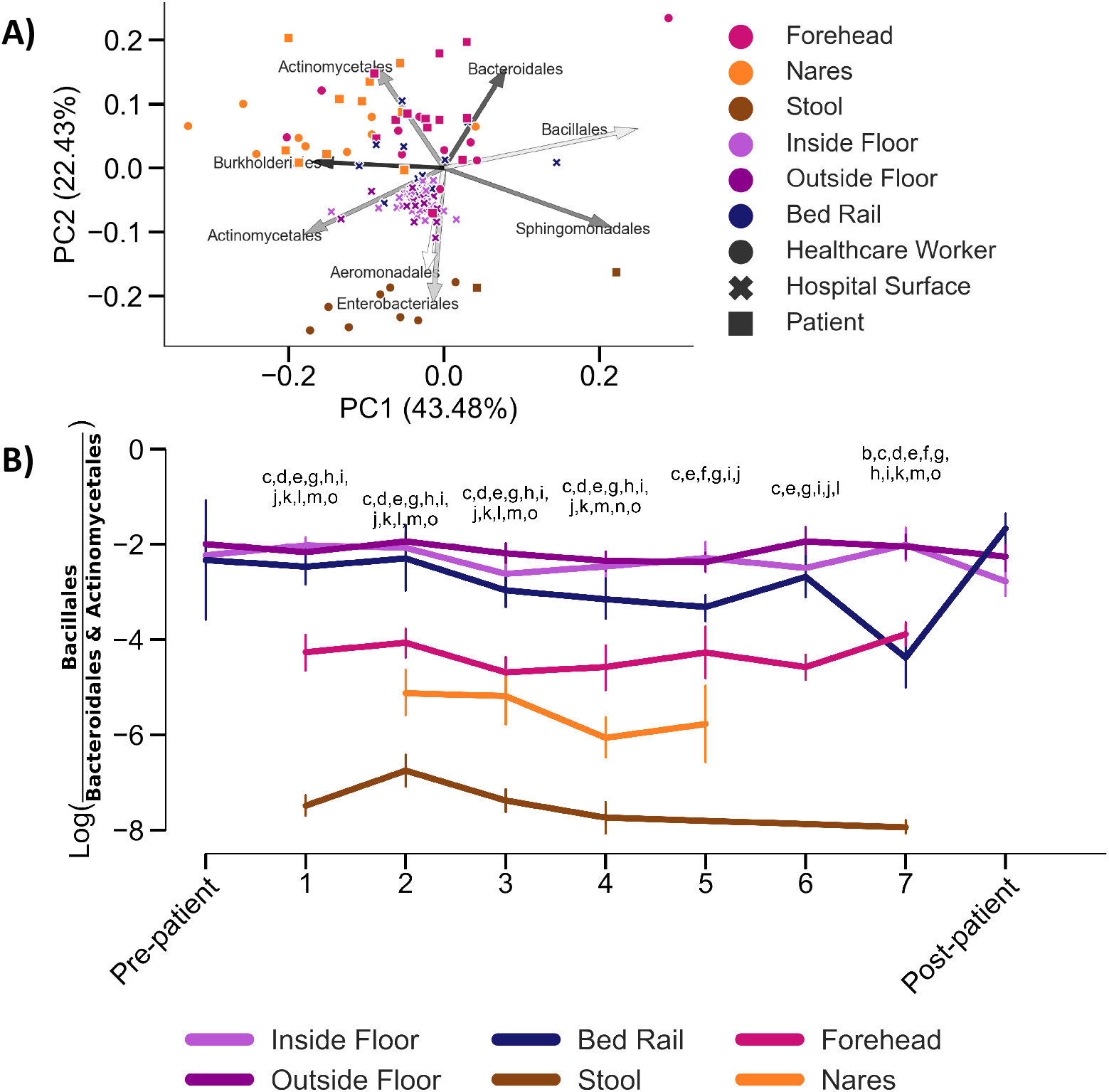
Longitudinal beta-diversity analyses of patients, health care workers and surfaces. **A**) Beta-diversity of human (n = 171; forehead, nares, and stool) and surface (n= 242; bed rail, inside and outside floor) samples accounting for repeated time point measures by Compositional Tensor Factorization (CTF). Arrows represent the top eight ASVs with the highest loadings, and are labelled by their order classification. **B**) Trajectory of differentially abundant taxa in human and surface samples across time. Lowercase letters represent pairwise comparisons with Bonferroni-corrected p-values <0.05; Inside Floor vs Outside Floor (a), Inside Floor vs Bed rail (b), Inside Floor vs Nares (c), Inside Floor vs Stool (d), Inside Floor vs Forehead (e), Outside Floor vs Bed rail (f),Outside Floor vs Nares (g),Outside Floor vs Stool (h),Outside Floor vs Forehead (i), Bed rail vs Nares (j), Bed rail vs Stool (k), Bed rail vs Forehead (l), Nares vs Stool (m), Nares vs Forehead (n), Stool vs Forehead (o). Full statistics in Data File S1.

### Positive association of microbial diversity and biomass with SARS-CoV-2

Next, we evaluated potential alpha diversity differences associated with SARS-CoV-2 detection. Overall, Faith’s phylogenetic alpha-diversity was significantly higher among surface samples than patient or health care worker samples (Fig. 4A). Across all sample types, Faith’s phylogenetic diversity tended to be higher in SARS-CoV-2 positive samples, and was significantly higher in forehead, inside floor, and outside floor samples (Fig 4B).

**Figure 4.**
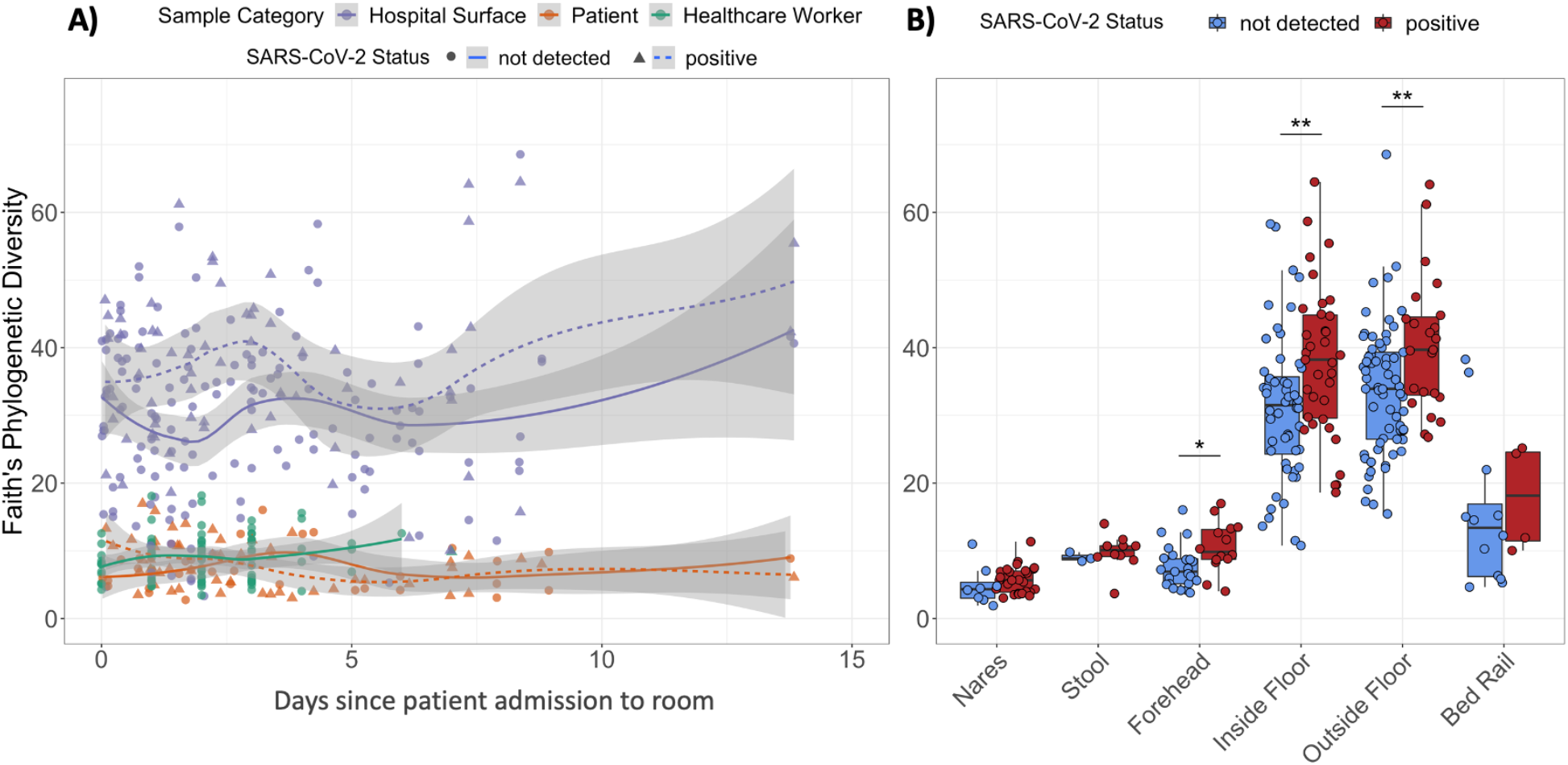
Alpha-diversity is higher in SARS-CoV-2 positive samples. **A**) Faith’s phylogenetic Diversity (rarefied to 4,000 reads per sample) of human and surface samples over time, fitted with locally estimated scatterplot smoothing (LOESS) curves. **B)** Faith’s phylogenetic diversity of humans and their surface samples grouped by SARS-CoV-2 screening results. Statistics resulted from Wilcoxon signed rank tests; *p<0.05, **p<0.01.

The high alpha-diversity of floor samples and significant association with SARS-CoV-2 detection led us to examine potential differences in biomass across floor samples. Two independent metrics were used to assess biomass; 16S rRNA gene amplicon sequencing read count, which because of our equal volume sequencing library pooling approach correlates with total bacterial load *(39, 40)*, and the Ct value from the CDC’s human RNAse P RT-qPCR target, which correlates with human biomass. 16S read count and human RNAse P Ct values are indirect measures of total bacterial and human biomass, respectively, and were significantly correlated (Pearson R^2^ = -0.40, p<0.0001). 16S read count was significantly higher in floor samples with detected SARS-CoV-2, but did not correlate with the number of viral copies detected (Fig. 5B). The abundance of human RNAse P was also significantly higher in floor samples with SARS-CoV-2 (lower Ct values), and positively correlated with viral load (Pearson R^2^ = -0.31, p-value = 0.011) (Fig. 5C); this correlation was not observed for the other sample types examined (nares, forehead, stool, bed rail). These results suggest that due to gravity SARS-CoV-2 is more likely to be detected on floors with high load of total microbial and human biomass.

**Figure 5.**
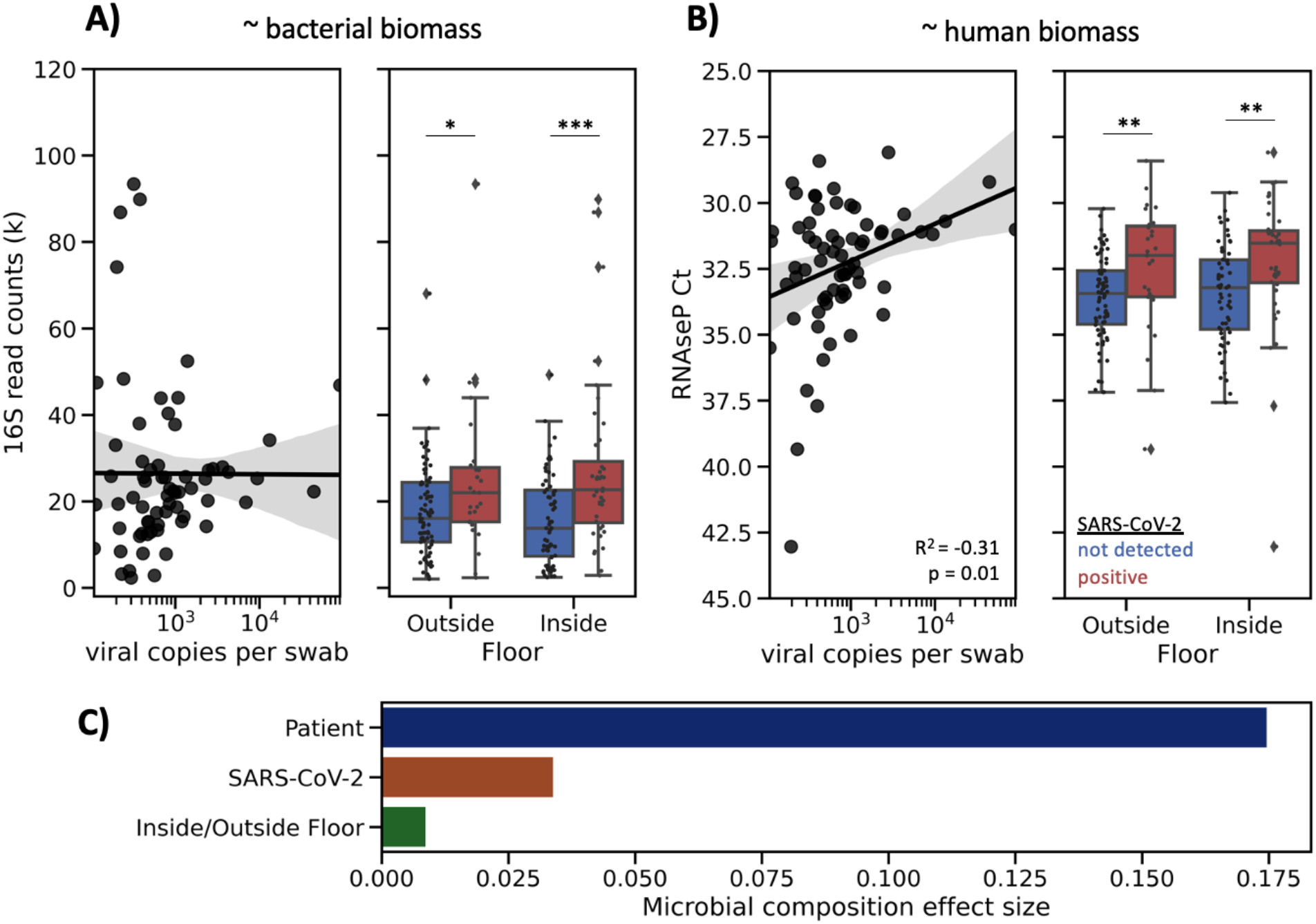
Floor sample SARS-CoV-2 status is associated with higher biomass and significantly contributes to microbial composition. (**A**) Abundance of 16S rRNA gene amplicon sequencing read count in SARS-CoV-2 positive floor samples showing no correlation with SARS-CoV-2 viral load. (**B**) Ct value of human RNAse P in SARS-CoV-2 positive floor samples showing significant correlation with SARS-CoV-2 viral load. Statistical analysis of scatter plots represents Pearson correlation, and box plots represents independent t-tests; *p<0.05, **p<0.01, ***p<0.001. (**C**) Effect size of significant, non-redundant variables identified from Redundancy Analysis on unweighted UniFrac PCoA of floor samples.

To determine if SARS-CoV-2 affected microbial composition in the built environment, we performed forward stepwise redundancy analysis *(41)* on unweighted UniFrac *(42)* principal components from floor samples (n=215). We chose floor samples for this analysis since floor samples had the largest number and highest biomass of all surfaces sampled (Fig. S6). Three non-redundant variables had a significant effect size, explaining a total of 21.7% variation in the data (Fig. 5C). The variable with the strongest effect size was patient identity (17.5%, p-value = 0.0002), which aligns with previous work demonstrating that the built environment microbiome is contributed from the humans inhabiting that space *(21)*. Whether the sample was an inside floor sample (next to patient bed) or outside floor sample (hallway directly in front of patient room) also had a small, yet significant effect size (0.8%, p-value=0.04). Importantly, SARS-CoV-2 detection status also significantly contributed to microbial variation (3.4%, p-value = 0.0004).

### Unique microbial signatures predict SARS-CoV-2 across patient sample types

To identify microbial features associated with SARS-CoV-2 positive samples, we independently trained Random Forest (RF) classifiers on nares (N=76), stool (N=44), and forehead samples (n=79) from COVID-19 patients and health care workers. Based on 16S rRNA gene amplicon sequencing microbial profiles, the RF models predicted SARS-CoV-2 status (positive vs. not detected) with 0.89 area under the receiver operating characteristic curve (AUROC) in unseen nares samples (Fig. 6A). Strikingly, skin (AUROC = 0.79) and stool (AUROC = 0.82) also showed high classifier accuracy. As the SARS-CoV-2-negative samples were overrepresented in the data, we also employed the area under the precision recall curves (AUPRC) to evaluate the prediction performance of each classifier, which were 0.76, 0.72, and 0.7 for nares, stool and forehead, respectively (Fig. 5B). A RF model built from bacterial profiles on the inside floor also showed a moderate prediction accuracy for discriminating SARS-CoV-2 status (AUROC=0.71; AUPRC=0.6, Fig. 5A and B). RF classifiers trained on outside floor and bed rail samples did not perform well, especially in the precision recall curves (Fig. S7).

**Figure 6.**
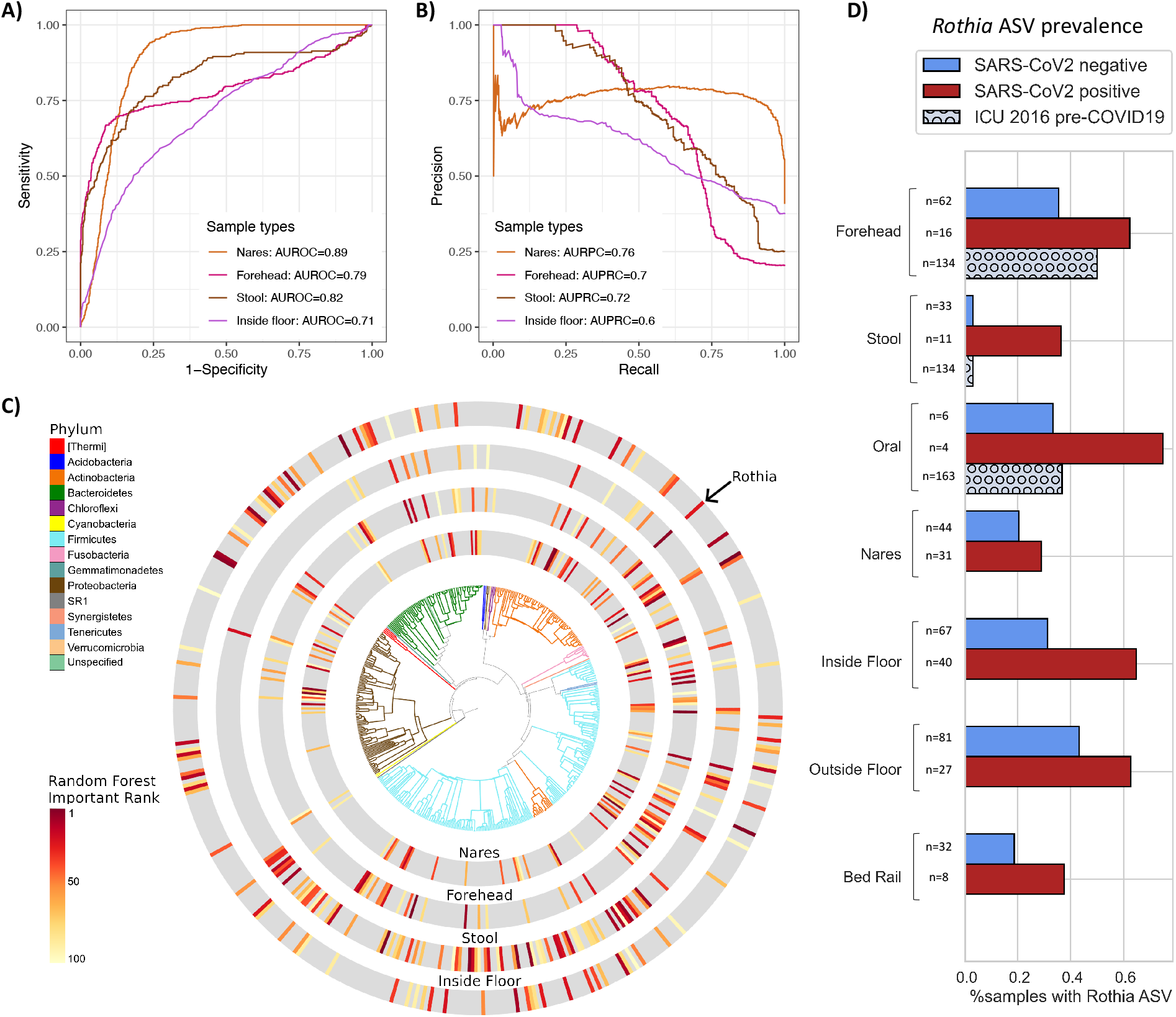
Bacterial composition is predictive of SARS-CoV-2 status in nares, forehead, stool and inside floor samples. The prediction performance of Random Forest classifiers on SARS-CoV-2 status for each sample type was assessed using AUROC (**A**) and AUPRC (**B**) for nares (n=76), forehead (n=79), stool (n=44), and inside floor (n=107), in a 100-fold cross-validation approach (see methods). (**C)** EMPress plot of the 100 features most predictive of SARS-CoV-2 status in nares, forehead, stool and inside floor samples, where a single ASV with 100% alignment to *Rothia dentocariosa* was identified across all sample types. Top 100 random forest importance ranks and GreenGenes taxonomy from nares, forehead, stool, and inside floor samples are available in Data File S2. (**D)** Proportion of samples containing the highly predictive *Rothia dentocariosa* ASV in SARS-CoV-2 positive and negative samples from the current study, and from *(30)* (ICU 2016 pre-COVID19).

The phylogenetic relationship of the top 100 ranked amplicon sequence variants (ASV) from the RF models were visualized with EMPress *(43)* (Fig. 5C). Stool and inside floor samples each had distinct sets of taxa driving the RF model compared to nares and forehead samples, which were more similar. Many of the highly ranked ASVs in the stool samples are from the class *Clostridiales*, a polyphyletic group of obligate anaerobes that were also identified as predictive of SARS-CoV-2 status in a wastewater study *(2)*.

ASVs from the genera *Actinomyces, Anaerococcus, Dialister, Gemella*, and *Schaalia* were in the top 40 ranked features of both forehead and nares samples (Data File S2); these taxa are normally found in anterior nares samples *(44–46)*, but are not commonly described in forehead microbiome samples. Interestingly, from Figure 2C, we observed that the unweighted UniFrac distance between samples from the same individual’s nares and forehead were more similar in COVID-positive room surfaces, suggesting that patients who shed virus into their environment could be cross-contaminating bacteria between nares and forehead (Fig. S8).

One ASV with an exact match to *Rothia dentocariosa* (GenBank ID <u>CP054018.1</u>) was highly ranked across all four disparate sample types: nares, forehead, stool, and inside floor. Further investigation shows this ASV is more prevalent in SARS-CoV-2 positive samples across all sample types examined. To exclude the possibility of this *Rothia* ASV being associated with sick patients generally, we examined the prevalence of this ASV in an intensive care unit microbiome study that was performed in 2016 *(30)*, and found that high *Rothia* prevalence is specific to SARS-CoV-2 positive patient samples (Fig. 5D).

## Discussion

The COVID-19 pandemic continues unabated as outbreaks ebb and flow around the globe. Because evidence for the synergistic effects of host-associated bacteria on viral pathogen stability and transmission continues to emerge, we set out to identify possible correlations between host- or surface-associated bacteria with SARS-CoV-2 presence and abundance in the built environment. At the onset of sampling, no hospital rooms or health care workers enrolled in the study had known exposure to SARS-CoV-2. Despite patients continually testing positive and shedding virus resulting in consistent surface contamination in the patient rooms, all samples collected from health care workers providing direct patient care to patients with COVID-19 were negative by both clinical RT-qPCR and antibody tests (data not shown). This includes the 3 health care workers who collected samples for the study. Aside from one stool sample where one of three viral targets amplified in our screening, all of the health care worker samples in this study (n=113) were negative for SARS-CoV-2, similar to findings from previous studies of exposed health care workers using airborne, contact and droplet protective PPE *(47–49)*. This contrasts with early reports of high SARS-CoV-2 transmission levels among health care workers before the implementation of general hospital-wide masking of healthcare workers and patients and of eye protection when interacting with an unmasked patient *(50, 51)*. Our findings highlight the importance of providing healthcare workers with appropriate PPE and with rigorous training in donning and doffing procedures to minimize self-contamination. In this hospital, the infection prevention measures (universal masking, eye protection, and appropriate PPE) were effective in preventing transmissions.

In this study, approximately 16% (83/529) of surface samples from hospital rooms occupied by COVID-19 patients and 6% (13/205) of surface samples from hospital rooms not currently occupied by COVID-19 patients had detectable levels of SARS-CoV-2. Not surprisingly, of the various surfaces sampled in this study, floor samples had the highest prevalence of SARS-CoV-2 detection. The intense and frequent oropharyngeal, respiratory, skin, bowel care provided to these critically ill patients is expected to produce shedding and contamination of the environment in close proximity of the patient, including the floors. Our findings replicate previous studies where floors had the highest prevalence of SARS-CoV-2 of all hospital room surfaces *(52, 53)*. Previous studies of environmental contamination report higher surface prevalence of SARS-CoV-2 in hospital settings, ranging from 25% to over 50% *(52, 54–56)*. The lower SARS-CoV-2 prevalence rates in this study could be due to differences in sampling strategy (e.g. area sampled, storage and extraction methods), more careful environmental cleaning of high touch areas around the patient, or due to physiological differences since different surface types differentially influence viral persistence *(57)*. Furthermore, contamination of hospital room surfaces with SARS-CoV-2 tends to be highest during the first 5 days after symptom onset (Chia et al., 2020). All patients enrolled in our study had symptoms for at least 6 days before admission to the hospital and enrollment in this study.

While SARS-CoV-2 was identified via RT-qPCR for both patient and hospital room samples, it cannot be determined whether the detected virus was viable. Infectivity is both a function of viral viability and abundance. One study assaying infectivity and RT-qPCR in parallel showed that samples with Ct values >30 were not infectious *(56)*. In our study, only 2 out of 79 positive surface samples amplified at least one SARS-CoV-2 target under 30 cycles, suggesting a relative low viral abundance. Interestingly, both of these samples were from the floor directly next to the patient bed in rooms that hosted patients who were mechanically ventilated during their stay. One of these potentially infectious samples was collected after the patient was transferred to the ICU and after room cleaning, and there were no other surface positives detected at that same time point. The other low-Ct floor sample came from a room where the patient had a consistently high viral load (Fig. S3B). However, the high Ct values for a majority of built environment samples in this study, and the lack of health care worker infection, suggest that the positive surfaces identified are an unlikely source of viral transmission in the hospital setting when contact precautions (gowns and gloves) are used correctly.

It should be acknowledged that transportation of samples in ethanol (to ensure the safety of those handling samples, as well as to enable microbiome analysis) instead of using viral transport media may have resulted in overall lower viral RNA yield. Despite these potential sources of variation, we found that bed rail and patient samples were highly similar in microbiomes to one another before cleaning, but this similarity disappeared after cleaning. Microbial community composition was also more similar between humans and the surfaces they touched (including between health care workers and keyboards, as well as patients and bed rails), supporting the robustness of our microbial sample collection and processing protocols.

It is both a strength and a limitation of this study that standard of care environmental cleaning was performed and was not influenced or altered by the study team. The daily cleaning regimen can vary depending on staff and other variables (hospital room surface types and disinfection protocols are summarized in Table S1) which is representative of hospital environmental practices worldwide. SARS-CoV-2 was amplified from floor samples, albeit at a relatively low abundance based on Ct values, in rooms even without COVID-19 patients and after cleaning. This highlights the importance of maintaining effective cleaning practices to mitigate the risk of viral spread via fomites. Although transmission risk from the floor is likely negligible as discussed above, the relatively high positivity rate for floor samples allowed us to use them as a proxy to study how microbial communities are interrelated with shed virus.

In the built environment, microbial load, human biomass and alpha-diversity were higher in floor samples positive for SARS-CoV-2. Floor samples also had the highest biomass of all the surface samples tested, including high-touch surfaces (e.g. bedrail, keyboard, door handles). This may help explain the higher prevalence of positive floor samples in COVID-19 patient rooms (39%) versus bed rail samples (11%), despite their distance from the patient. This is in agreement with previous research showing that bacterial- and viral load are positively correlated in built environment samples *(58)*. The relatively low prevalence of SARS-CoV-2 contamination on bed rail samples may also be because many of the patients were deeply sedated and were not actively moving in bed including touching the bedrails or because high touch areas in close proximity to the patient are cleaned by nurses at each shift, and/or due to differences in material (vinyl versus plastic).

Using Random Forest models to classify microbes associated with SARS-CoV-2 detection, we found 16S microbial profiles had high predictive accuracy of SARS-CoV-2 presence in nares, stool, forehead, and inside floor samples. Despite these sample types having distinct microbiomes covering a broad range of microbial diversity (Fig. 2), we identified a single *Rothia* ASV that was highly ranked in the Random Forest classifier across all four sample types. This ASV was also more prevalent in SARS-CoV-2 positive samples across all human sample types and floor and bed rail samples in our dataset. By comparing the prevalence of this ASV across our dataset and a 2016 study from an intensive care unit *(30)*, we found that this signal is specific to SARS-CoV-2 positive samples, and not other factors associated with an ICU admission such as antibiotic use. This finding supports previous work reporting *Rothia* to be enriched in SARS-CoV-2 positive stool *(59)* and bronchoalveolar lavage fluid *(60)*, and further suggests a role in nares, forehead, and surfaces.

While the mechanism remains unclear, the consistent *Rothia* ASV prevalence trend across both patient and surface sample types suggest an association of this bacteria with SARS-COV-2. Species from the genus *Rothia* are common to the human oral microbiome *(61)*, but have also been identified as opportunistic pathogens *(62)*. Oral microbes have been found to colonize the gastrointestinal tract, especially in disease states *(63)*. This suggests a possible increased oral-fecal transmission triggered under viral infection that manifests as a hallmark of COVID-19. Interestingly, we also found that patients with cardiovascular disease comorbidities tended to have higher prevalence of the *Rothia* ASV associated with SARS-CoV-2, compared to patients with pre-existing cardiovascular disease (45% versus 26%, respectively). *Rothia dentocariosa* can cause endocarditis, particularly in patients with a history of cardiovascular disease *(62, 64)*. Using data from the American Gut Project *(65)*, we tested for the presence of this *Rothia* ASV in samples from those self reporting a medical diagnosis of a cardiovascular disease, and those self reporting not having a cardiovascular disease. We observed a significantly higher prevalence of *Rothia* in samples with a medical reporting (Fisher’s exact test, p=0.041) than those without, suggesting that *Rothia* may be associated with cardiovascular disease even outside of the context of SARS-CoV-2. Cardiovascular disease can predispose individuals to worse outcomes with COVID-19, and COVID-19 itself can cause cardiovascular problems *(66)*. Further studies are required to determine the mechanism underlying this association and how it may be translated into effective methods for reducing SARS-CoV-2 transmission.

This large-scale study is the first to examine the microbial context of SARS-CoV-2 in a hospital setting. We detected viral contamination across a variety of surfaces in the ICU and the general medical-surgical unit, including rooms that were not used to treat patients with COVID-19 infection. Nonetheless, current hospital infection prevention measures including standard environmental cleaning and the use of PPE were adequate in preventing hospital transmission of SARS-CoV-2 to healthcare workers who directly provided care to patients with COVID-19 infection. Across a remarkable diversity of microbiomes (floor, nares, stool, skin), we identified a single bacterial ASV, *Rothia dentocariosa*, that was highly predictive of and co-identified with SARS-CoV-2. This association could be a result of direct interactions with the virus, or indirect correlations through effects on the host, but both possibilities present exciting new avenues to combat SARS-CoV-2 virulence. Our discovery of bacterial associations with SARS-CoV-2 both in humans and the built environment demonstrates that bacteria-virus synergy likely plays a role in the COVID-19 pandemic.

## Materials and Methods

### Study Design

#### Sample collection

Patients admitted to the UCSD Medical Center - Hillcrest who were either confirmed COVID-19 patients or Persons Under Investigation (PUI: have symptoms and undergoing testing) were approached for informed consent upon admission. Patients whose clinical test was negative were included in the study as controls for surface sampling. Health care workers providing direct care for PUI’s and COVID-19 patients were included in the study. Following hospital policy, all underwent daily symptomatic screening and wore the following PPE during treatment of PUI and COVID-19 patients: goggles or face-shield, N95 mask, gown, gloves; hair and shoe coverings were available but inconsistently used. All participants were consented under UCSD Human Research Protections Program protocol 200613.

We followed the excretion pattern of the virus from the skin, respiratory tract, and gastrointestinal tract. From patients and health care workers, specimen samples were obtained from the forehead, nares, and stool. Additional throat swabs and/or tracheal aspirate samples were collected for a subset of patients and health care workers; ‘oral’ samples. Patient samples were collected by gloved health care workers via dual-tipped synthetic swabs which were immediately transferred to tubes containing 95% ethanol. Stool was collected from patient bed pans or from collection bags that were connected to a rectal tube. Health care workers self-collected swabs over a time series of 4 days. A chronological series was also employed for patient samples, with the target sampling schemes as follows: samples collected within the first 12 hours of hospital admission with sequential samples obtained once daily for the first 4 days of hospitalization and a subset of samples collected regularly until the patient vacated the room (Fig. 1A). Actual sample collection timing varied by patient availability and duration in the hospital (Fig. S3).

Dual-tipped polyester swabs (BD BBL CultureSwabs #220145) were pre-moistened by dipping for 5 seconds into 95% spectrophotometric-grade ethanol solution (Sigma-Aldrich #493511), then used to vigorously swab surfaces that are frequently in contact with health care workers or patients. Surfaces were swabbed for 10-15 seconds with moderate pressure, and swabs were returned to the collection container. Outside of patient rooms, prior to entering the room, the floor (1 foot at the entrance from the door) and outside door handle were swabbed. Inside patient rooms, the inside door handle, floor (1 foot near the patient’s bed on side closest to door), bedrail (side closest to door), and keyboard were swabbed. Depending on the patient room, if an air filter was present, the intake was swabbed. For a subset of samples, patient care equipment such as portable ultrasound and ventilator screen were also swabbed, as well as the toilet seat. After sample collection, dual-tipped swabs were returned to the swab container. Surface samples were collected at the same time as patient sample collection, as well as prior to patient admission and following patient discharge and room cleaning, when possible.

#### Nucleic acid extraction

Sample plating and extractions of all clinical and environmental specimens were carried out in a biosafety cabinet Class II in a BSL2+ facility. Sample swabs were plated into a bead plate from the 96 MagMAX™ Microbiome Ultra Nucleic Acid Isolation Kit (A42357 Thermo Fisher Scientific, USA). Following the KatharoSeq low biomass protocol (Minich 2018), each sample processing plate included eight positive controls consisting of 10-fold serial dilutions of the ZymoBIOMICS™ Microbial Community Standard (D6300 Zymo, USA) ranging from 5 to 50 million cells per extraction. Each plate also contained a minimum of 8 negative controls. Nucleic acids purification was performed on the KingFisher Flex™ robots (Thermo Fisher Scientific, USA) using the MagMAX™ Microbiome Ultra Nucleic Acid Isolation Kit (Applied Biosystems™), as instructed by the manufacturer. Briefly, 800 μL of lysis buffer was added to each well on the sample processing plate, and briefly centrifuged to bring all beads to the bottom of the plate. Sample swab heads were added to the lysis buffer and firmly sealed first with MicroAmp™ clear adhesive film (Thermo Fisher Scientific, UK) using a seal roller, and the sealing process repeated twice using foil seals. The plate was beaten in a TissueLyser II (Qiagen, Germany) at 30 Hz for 2 minutes and subsequently centrifuged at 3700 x g for 5 minutes. Lysates (450 μL/well) were transferred into a Deep Well Plate (96 well, Thermo Fisher Scientific, USA) containing 520 μL of MagMax™ binding bead solution and transferred to the KingFisher Flex™ for nucleic acid purification using the MagMax™ protocol. Nucleic acids were eluted in 100 µL nuclease free water and used for downstream SARS-CoV-2 real time RT-qPCR.

#### SARS-CoV-2 RT-qPCR and viral load quantification

The Center for Disease Control (CDC) 2019-Novel Coronavirus Real-Time RT-PCR Diagnostic Panel *(67)*, and the E-gene primer/probe from the World Health Organization *(68)*, were used to assess SARS-CoV-2 status via reverse transcription, quantitative polymerase chain reaction (RT-qPCR). Accordingly, each plate of extracted nucleic acid (96-well plate) was aliquoted into a 384-well plate with four separate reactions per sample; two reactions targeted the SARS-CoV-2 nucleocapsid gene (CDC N1 and N2), one reaction targeted the SARS-CoV-2 virporin forming E-gene (WHO E-gene), and one reaction targeted the human RNAse P gene as a positive control for sample collection and nucleic acid extraction (CDC).

Each reaction contained 3 μL of TaqPath™ 1-Step RT-qPCR Master Mix (Thermo Fisher Scientific, USA), 400 nm forward and reverse primers and 200 nm FAM-probes (IDT, USA - table with sequences below), 4 µL RNA template, and H2O to a final volume of 10 µL. Master mix and sample plating were performed using an EpMotion automated liquid handler (Eppendorf, Germany). Each plate contained both positive and negative controls. The positive control was vRNA and eight serial dilutions of viral amplicons for viral load quantification (details below). Six extraction blanks and one RT-qPCR blank (nuclease-free H_2_O) were included per plate as negative controls. RT-qPCR was performed on the CFX384 Real-Time System (BIO-RAD). Cycling conditions were reverse transcription at 50°C for 15 minutes, enzyme activation at 95°C for 2 minutes, followed by 45 cycles of PCR amplification (Denaturing at 95°C for 10 s; Annealing/Extending at 55°C for 30 s). Cycle threshold (Ct) values were generated using the CFX384 Real-Time System (BIO-RAD) software.

Viral load quantification was performed using a standard ladder comprising serially diluted target amplicons. SARS-CoV-2 viral RNA was reverse transcribed into cDNA using the Superscript IV enzyme (Thermo Fisher, USA) and PCR amplified with KAPA SYBR® FAST qPCR Master Mix (KAPA Biosystems, USA) using the N1, N2, and E gene primers in duplicate 20 µL reactions with cycling parameters as detailed above. Each amplicon reaction was run across a 1.5% agarose gel and the resulting bands were excised and purified into 100 µl nuclease-free water with the MinElute Gel Extraction Kit (Qiagen, Germany). Amplicons were quantified with in duplicate with the Qubit™ dsDNA HS Assay Kit (Thermo Fisher, USA) and copies per µL were calculated based on predicted amplicon length (N1 72 bp, N2 67 bp, and E gene 113 bp). Eight, 10-fold serial dilutions were added to the RT-qPCR for final estimated copy input per reaction of 10 million to one. Viral load per swab head was calculated by first using the slope and intercept from the N1 amplicon ladder linear regression per plate to determine the number of viral copies per reaction, and then multiplying this number by 25 since 4 µL out of a total 100 µL extracted nucleic acid was used as input to the RT-qPCR.

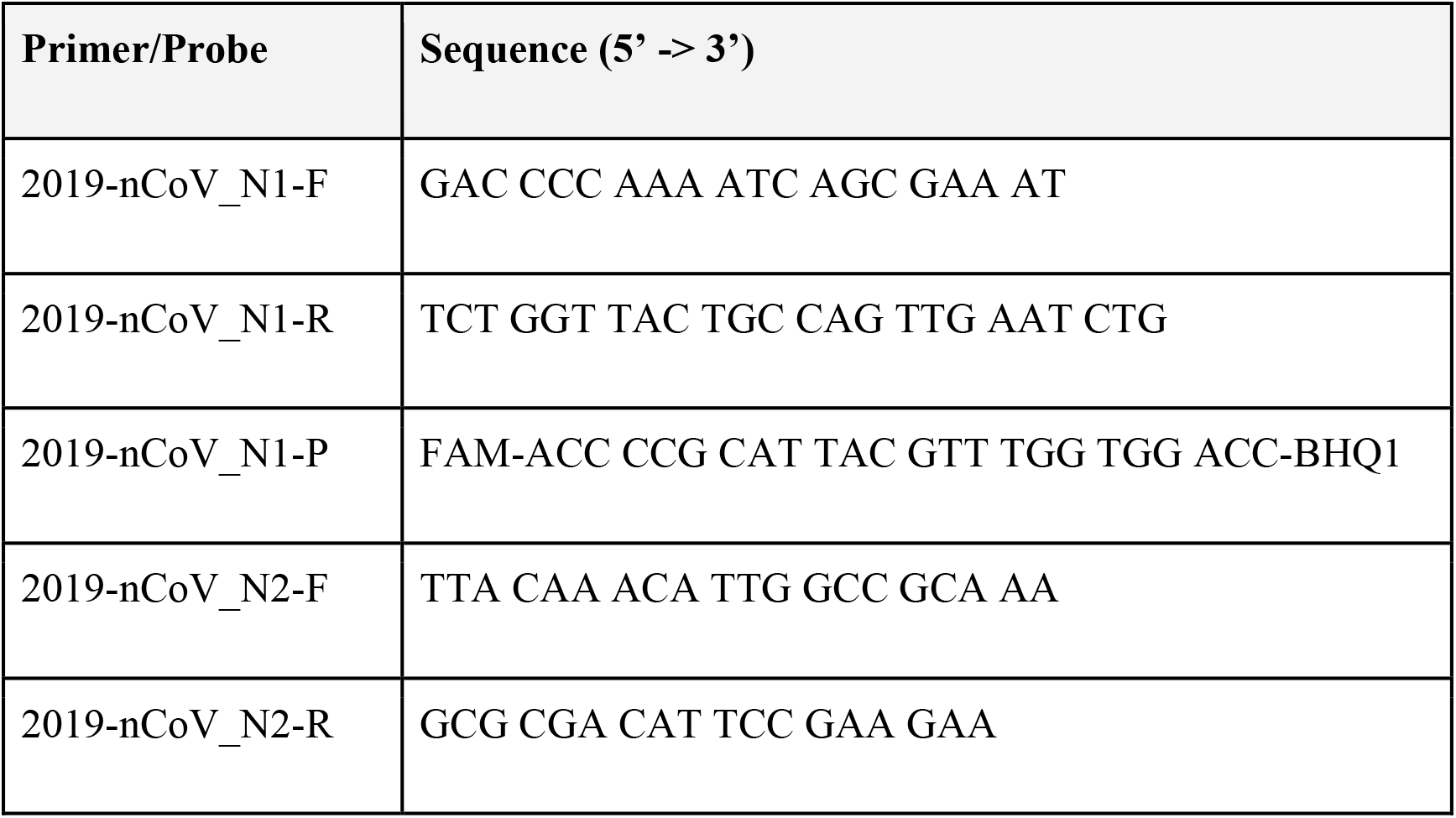
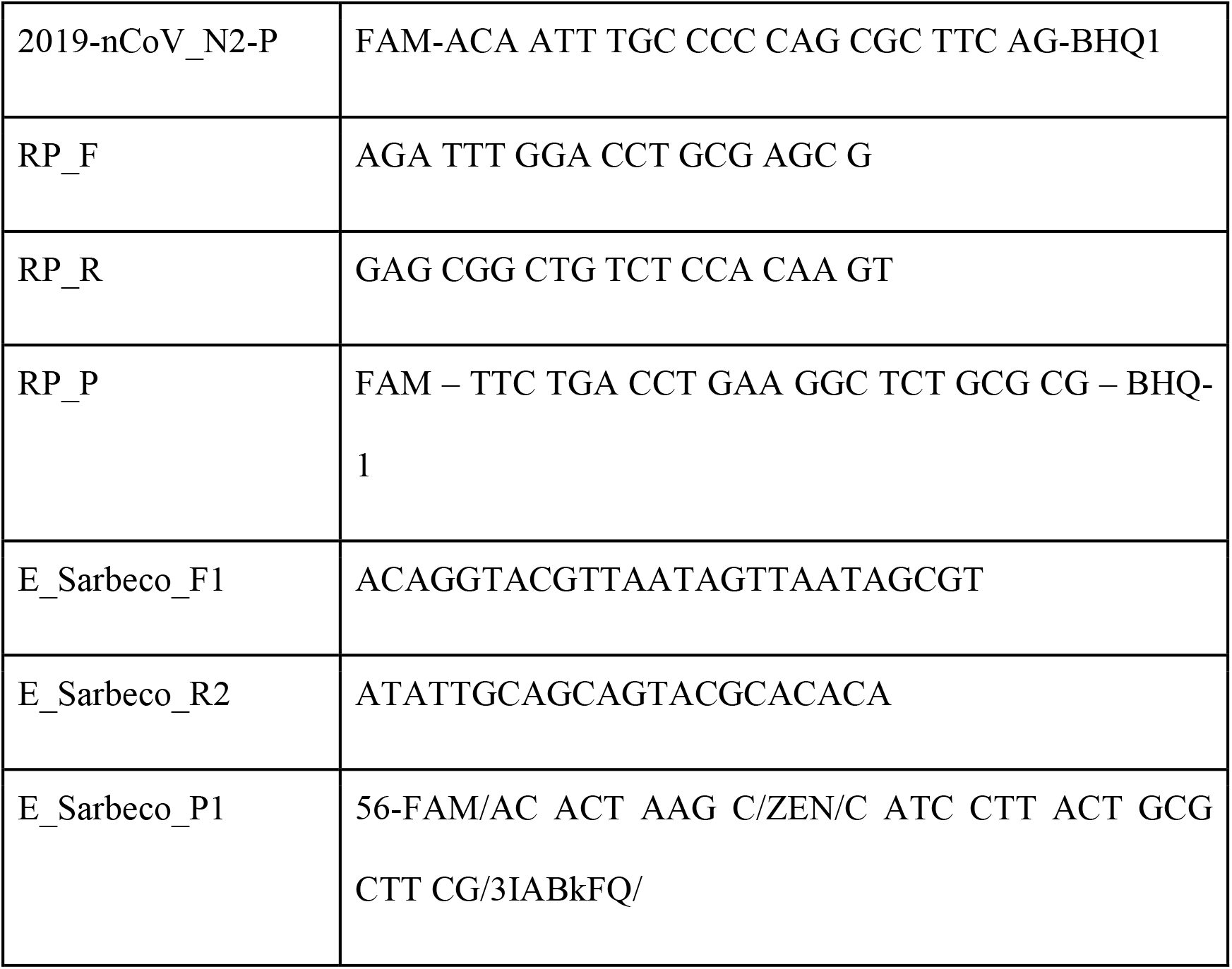

#### 16S rRNA gene amplicon sequencing

16S rRNA gene amplification was performed according to the Earth Microbiome Project protocol (Thompson et al., 2017). Briefly, Illumina primers with unique reverse primer barcodes (Caporaso et al., 2012) were used to amplify the V4 region of the 16S rRNA gene (515f-806rB, Walters et al., 2016). Amplification was performed in a miniaturized volume *(69)*, with single reactions per sample *(70)*. Equal volumes of each amplicon were pooled, and the library was sequenced on the Illumina MiSeq sequencing platform with a MiSeq Reagent Kit v2 and paired-end 150 bp cycles. Raw data is available through EBI under accession ERP124721 and associated feature tables are publicly available in Qiita (qiita.ucsd.edu) (Gonzalez et al., 2018) under study ID 13092.

### Statistical Analysis

#### Data pre-processing

Raw 16S rRNA gene amplicon sequencing data was demultiplexed, quality filtered, and denoised with deblur *(71)* through Qiita *(72)* under study ID 13092. Downstream data processing was performed using Qiime2 *(33)*. The serially diluted mock communities included in each extraction plate (see *Nucleic Acid Extraction* section) were used to identify the read count threshold at which 80% of sequencing reads aligned to the positive control according to the KatharoSeq protocol *(40)* (code available at https://github.com/lisa55asil/KatharoSeq_ipynb), and all samples falling below the threshold set for each independent sequencing run were removed from downstream analysis. The KatharoSeq-filtered feature tables were merged, and features present in less than three samples were removed from downstream analysis, with the final feature table containing 589 samples and 9461 features.

#### Beta-diversity analyses

To verify that study samples of particular types clustered with similar types from other microbial studies, we estimated the UniFrac phylogenetic distance between samples and visualized the distance of variation of our current project in reference to samples from the Earth Microbiome Project. For significance testing based on distances from sequencing data, a permutation test was used. This was chosen since univariate statistical tests often assume that observations are independently and identically distributed, which is not the case with distance calculations. Similar to PERMANOVA, the group labels were shuffled, and a Kruskal-Wallis test was applied. P-values were calculated by (#(K > Kp) + 1) / (number of permutations + 1) where K is the kruskal-wallis statistic on the original statistic and Kp is the Kruskal-Wallis statistic computed from the permuted grouping. 1000 permutations were used for the permutation test.

#### Longitudinal data analysis

To detect microbial changes over time without being limited by interindividual variation, we used a dimensionality reduction tool, compositional tensor factorization (CTF) *(34)*. This tool incorporates microbiome information from an individual host or sample source, which has been sampled across multiple time-points and reveals the net differences in microbial beta-diversity across sample types or patient profiles. We used Bayesian Sparse Functional Principal Components Analysis (SFPCA) *(73)* methodology to model temporal variations and sample type differences in viral load.

To quantify the contribution of potential source environments (i.e. patient microbiome) to the hospital surface microbiome (as a sink), SourceTracker2 *(31)* was used.

#### Random Forest Analysis

We performed machine learning analysis of bacterial profiles derived from 16S rRNA gene amplicon sequencing from multiple sample types (nares, skin, stool, inside floor, outside floor, and bed rail) to predict the samples’ SARS-CoV-2 status according to RT-qPCR (i.e., “positive” or “not detected”). For each sample type, a Random Forest sample classifier was trained based on the ASV-level bacterial profiles with tuned hyperparameters as 20-time repeated, stratified 5-fold cross-validation using the R caret package *(74)*. The dataset of each sample type was repeatedly split into five groups with similar class distributions, and we trained the classifier on 80% of the data, and made predictions on the remaining 20% of the data in each fold iteration. We evaluated each classifier using both area under the receiver operating characteristic curve (AUROC) and area under the precision-recall curve (AUPRC) based on the samples’ predictions in the holdout test set using the R PRROC package *(75)*. For all four sample types, our data had an imbalanced representation of SARS-CoV-2 status, and “not detected” was consistently the majority class (nares: 45 not detected vs. 31 positives; forehead skin: 63 not detected vs. 16 positives; stool: 33 not detected vs. 11 positives; inside floor: 67 not detected vs. 40 positive; inside floor: 81 not detected vs. 27 positives; bed rail: 38 not detected vs. 8 positives). To assess how well a classifier can predict the SARS-CoV-2 positive samples (the minority class) using microbiome data, the AUPRC was calculated by assigning “positive” as the positive class. Next, the importance of each ASV for the prediction performance of the four classifiers (for nares, forehead skin, stool, and inside floor) was estimated by the built-in Random Forest scores in the 100-fold cross-validation. For each body site or environmental site, we finally ranked all ASVs by their average ranking of importance scores in the 100 classification models. The code for generating the multi-dataset machine learning analysis is available at https://github.com/shihuang047/crossRanger and is based on Random Forest implementation from R ranger package *(76)*.

To identify the ASVs consistently important to the prediction of SARS-CoV-2 across the four different sample types, we visualized the top 100 ranked important ASV’s and their phylogenetic relationship for each sample type using EMPress *(43)*.

#### Redundancy Analysis

To quantify the effect size of different metadata variables on our 16S rRNA gene amplicon sequencing dataset, we applied redundancy analysis on the robust Aitchison principal coordinates analysis biplot *(77)* as described previously *(41)*. Briefly, RDA employs the *varpart* function in R which uses linear constrained ordination to estimate the independent and shared contributions of multiple covariates on microbiome composition variation.

## Data Availability

Sequencing data is available through the European Bioinformatics Institute under accession ERP124721.

https://www.ebi.ac.uk/ena/browser/view/PRJEB41002

## Acknowledgements

This work would not have been possible without the support of Louis-Felix Nothias, Chris Callewaert, and Alison Vrbanac, who transported samples from the hospital to the lab, Franck Lejzerowicz, Dom Nguyen, Emily Kunselman, Kanwal Aziz, and Megan Preovolos, who assisted with kit preparation, and Karsten Zengler who provided access to lab space. We are very thankful to the patients and health care workers who participated in the study.

## Funding

This work was partially supported by IBM Research AI through the AI Horizons Network and the UC San Diego Center for Microbiome Innovation (to SH, KC, YVB, and RK). RED is supported by NIH/NIGMS IRACDA K12 (GM068524) and the National Science Foundation PRFB (P2011025). RK is supported by NIH Pioneer (1DP1AT010885) and NIH/NIDDK (1P30DK120515).

## Author contributions

S.M.A, F.A., J.A.G, R.K., and D.A.S. conceived of the study. C.M., P.B.-F., S.M.A., D.A.S., and F.A. developed the sample collection and processing methodology, and F.A., L.C., and D.A.S collected samples and metadata. Y.V.-B., S.M.A., and G.A. curated metadata. C.M., P.B.-F., S.K., S.D., G.E.-M., N.G., M.C.S.G., M.B., K.S., and G.H. processed samples. C.M., P.B.-F., P.D., S.H., K.C., L.J., C.M., R.E.D., G.R., D.M., G.A., R.S., J.P.S., and S.M.A conducted formal analysis and visualization. N.H., K.L.B., H.-C.K., A.P.C., L.P, and Y.V.-B. supervised and provided feedback on formal analysis and visualization. F.J.T. and D.A.S. provided a clinical perspective to interpretation of results. C.M., P.B.-F., and S.M.A. wrote the original manuscript, and all authors edited and approved the final manuscript.

## Competing interests

The authors have no competing interest to declare.

## Data and materials availability

Sequencing data is available through the European Bioinformatics Institute under accession ERP124721. Additionally, sequencing data and processed tables and taxonomy assignments are available through QIITA *(72)* under study ID 13092.

## Supplementary Materials

**Figure S1.**
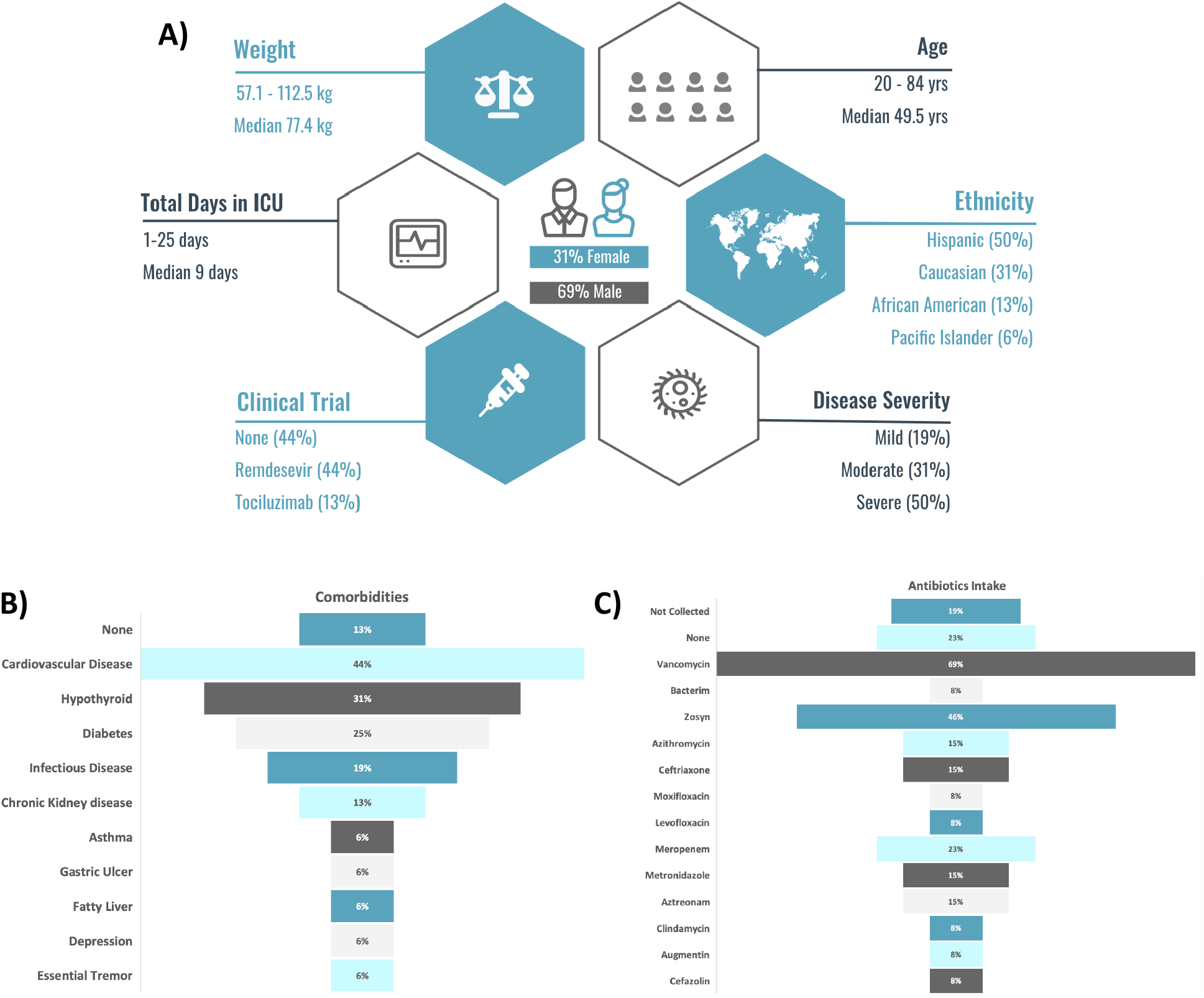
Patient (n=16) demographics (A), antibiotics intake (B), comorbidities (C).

**Figure S2.**
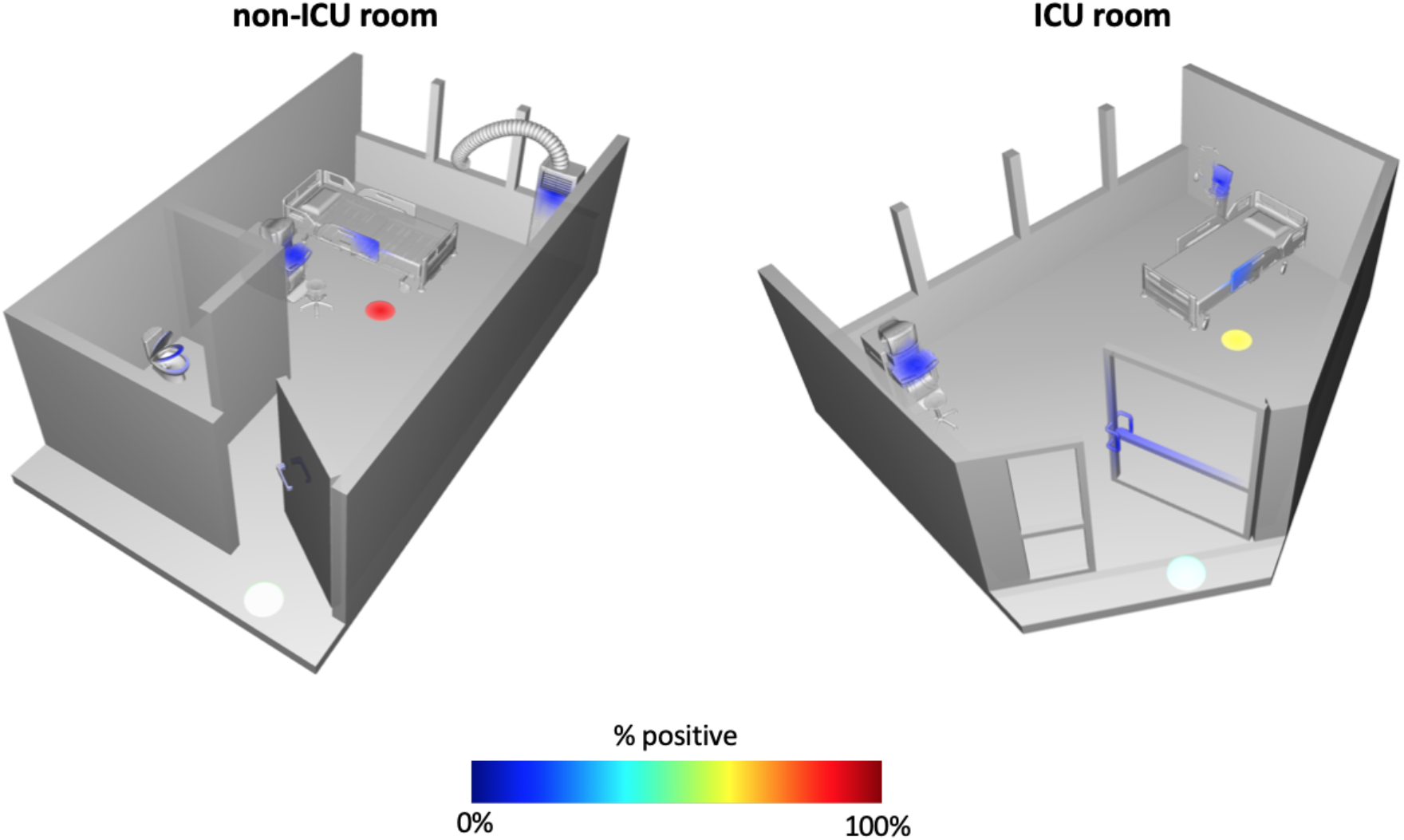
Ili’ spatial mapping of standard hospital (non-ICU) room and intensive care unit (ICU) room. Heatmap depicts the percent of samples collected at each site that were positive for SARS-CoV-2.

**Figure S3.**
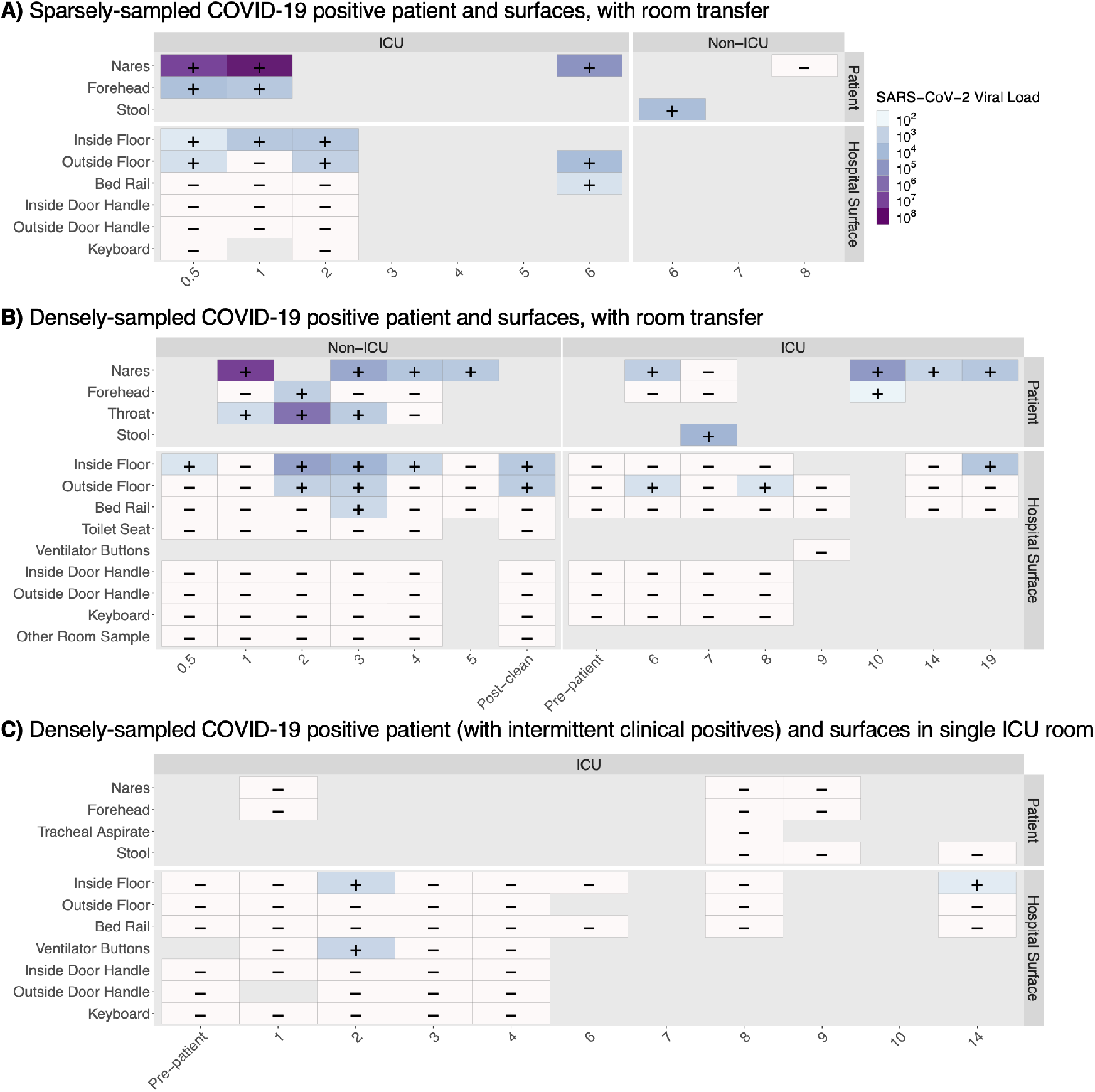
Snapshot of variability in longitudinal sample collection and SARS-CoV-2 viral load per swab between patients and their hospital rooms, starting at patient admission time. For samples where SARS-CoV-2 was detected (+), a darker color indicates a higher viral load. White boxes represent samples with no detectable virus (-). Patient **A** was admitted 12 days after symptom onset and was moved to a general surgery unit room after 6 days in the ICU. Patient **B** was admitted 8 days after symptom onset and moved from general surgery to the ICU, where they were intubated. Patient **C** was admitted to the ICU 9 days after symptom onset, and despite having symptoms consistent with COVID-19 repeatedly tested negative by clinical nasopharyngeal swab; their only clinical positive came from a tracheal aspirate sample mid-way through their stay in the ICU.

**Figure S4.**
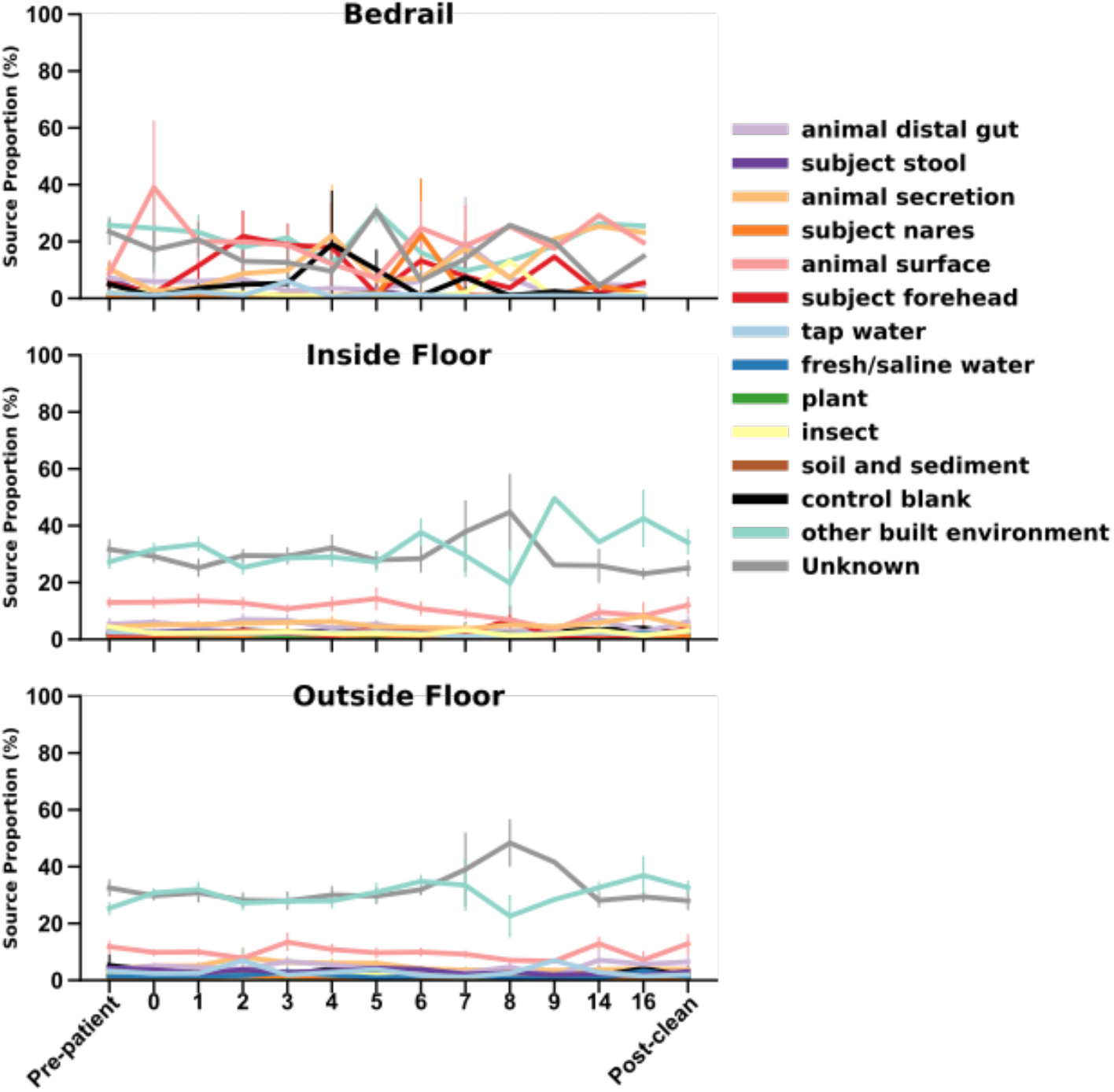
Source tracker on meta-analysis data. Floor samples formed a distinct cluster in this dataset; source tracking *(31)* with floor samples (n=215) as the sink and meta-analysis samples (n=1,990) as the source reveals that these floor samples match other built environment samples. The other built environment samples included in this meta-analysis were mostly floor (27.7%), faucet handles (19.6%), and gloves (15%).

**Figure S5.**
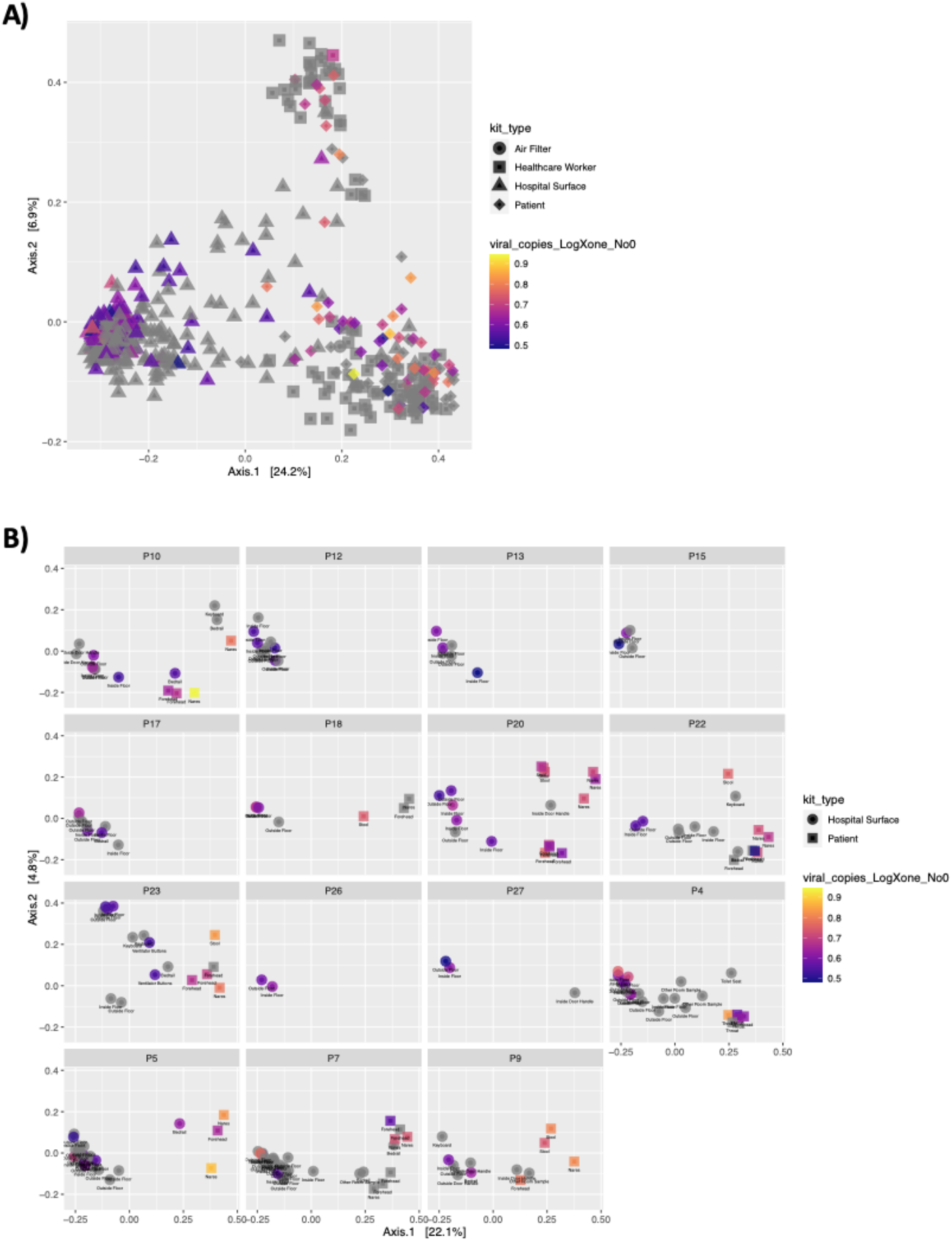
Beta-diversity has a statistically significant but weak correlation with viral load. PCoA of unweighted UniFrac distances between samples, with SARS-CoV-2 positive samples colored by viral load across the whole dataset (**A**) and subset by each patient with at least one surface positive (**B**). Statistical analysis performed with Adonis (PERMANOVA) found a small (R^2^ < 0.01) but significant (*p-value* = 0.043) association between beta-diversity and viral load across all samples.

**Figure S6.**
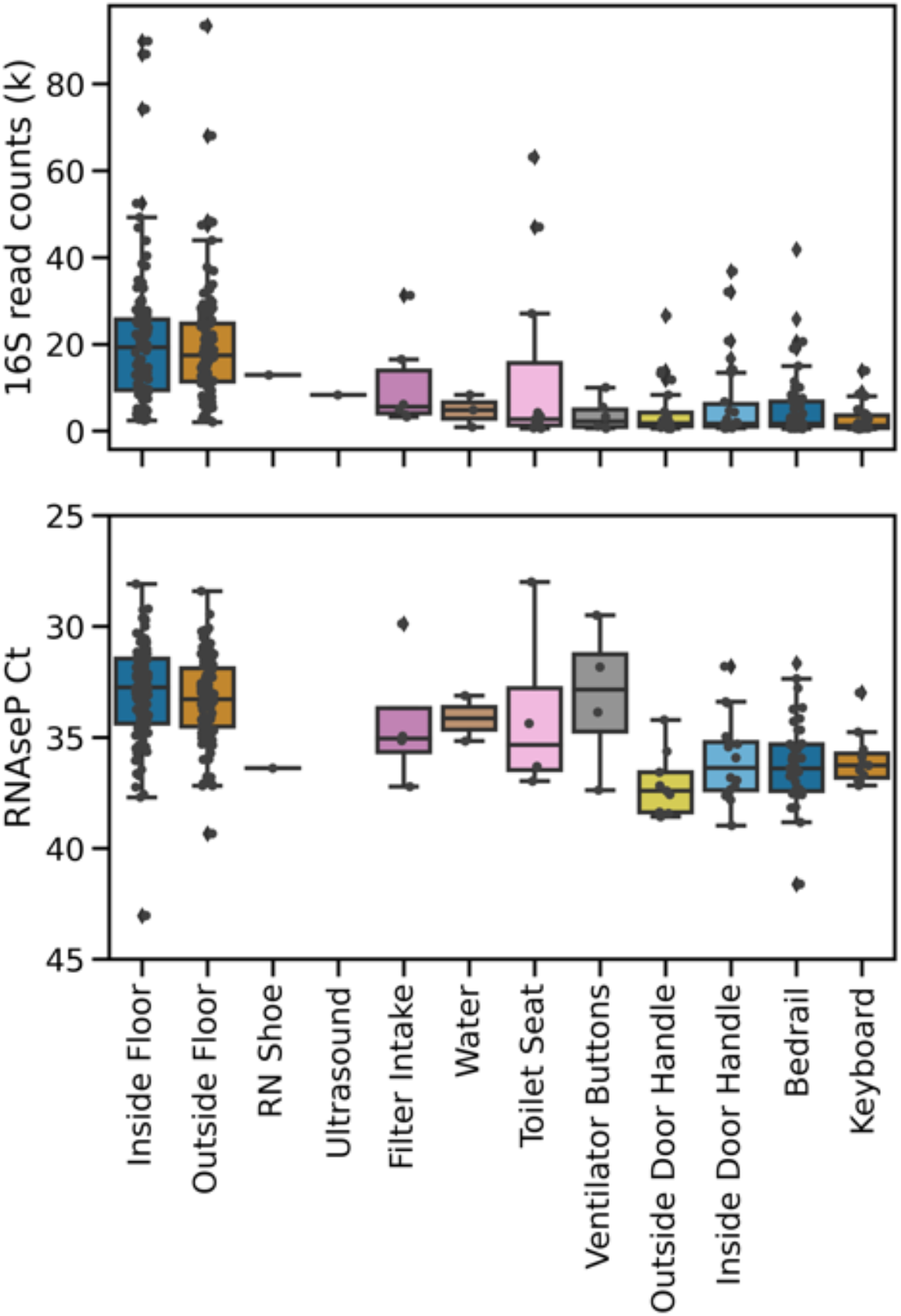
Bacterial (16S rRNA gene amplicon sequencing read count) and human biomass (RNAse P Ct) is higher in floor samples than other surface sample types.

**Figure S7.**
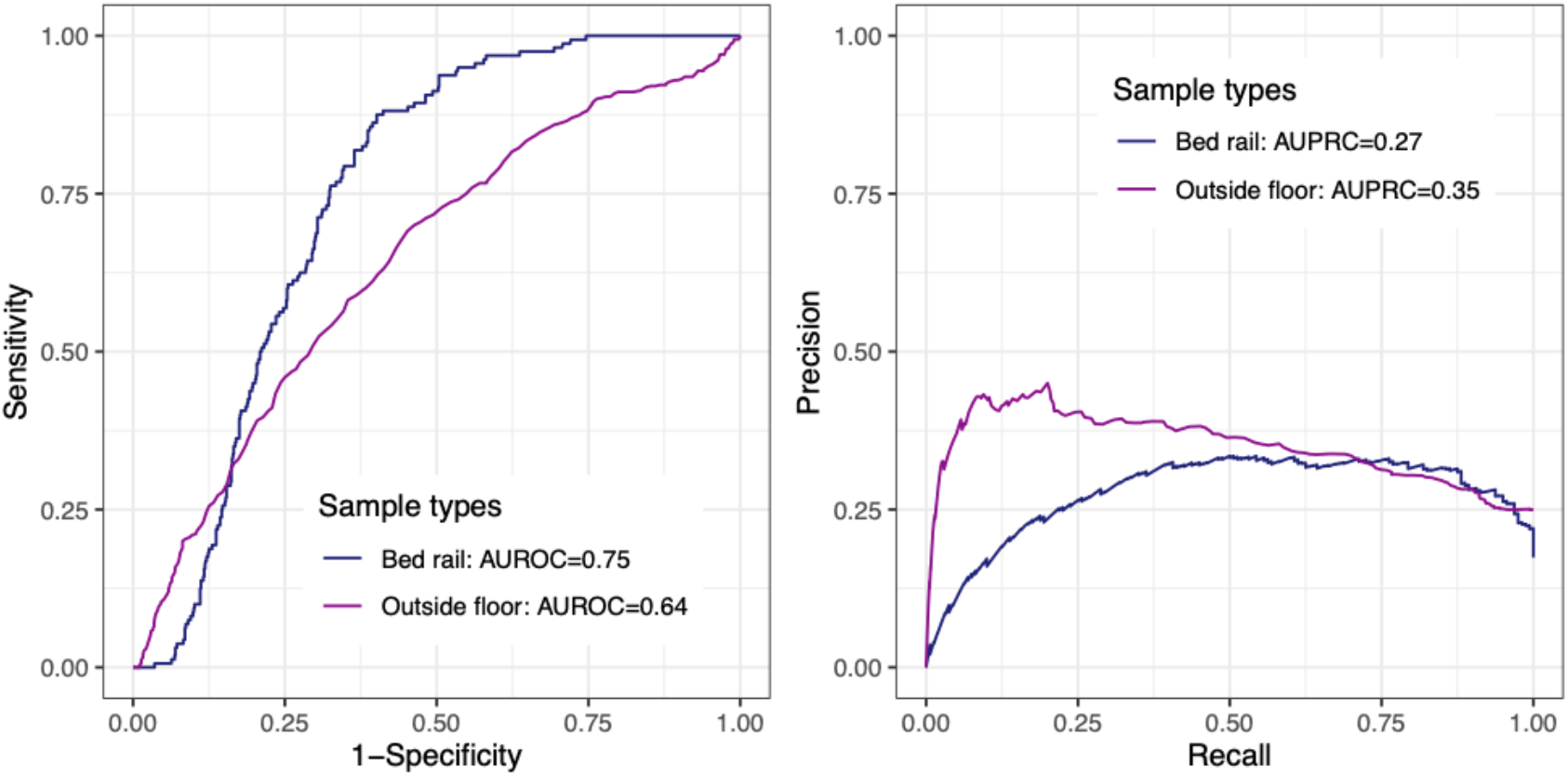
Random Forest classifier performance with 100-fold cross validation in the outside floor (n=108; 81 not detected vs. 27 positives) and bed rail samples (n=46; 38 not detected vs. 8 positives).

**Figure S8.**
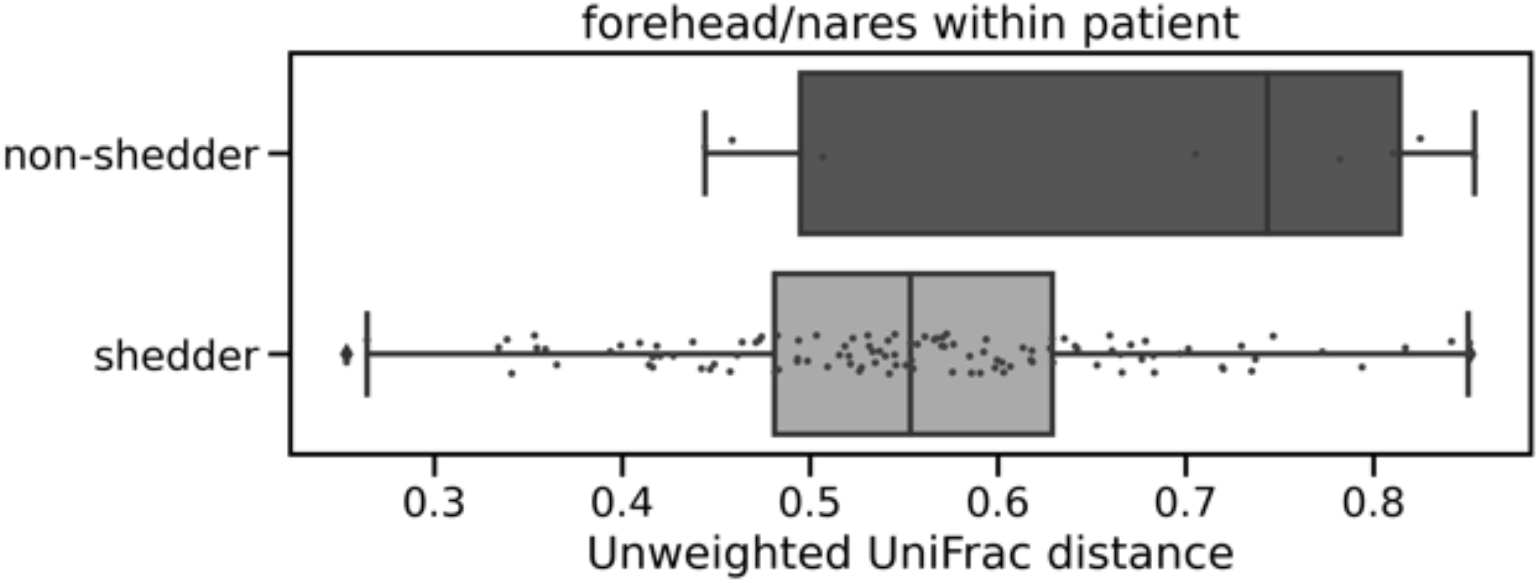
Unweighted UniFrac distance between forehead and nares samples from the same host. ‘Shedder’ (n=12) is a patient who had detectable virus on the surface in their room and ‘non-shedder’ (n=4) did not. Bootstrapped Kruskal-Wallis p-value is 0.003.

**Table S1.** Hospital surface materials and cleaning practices.

| Surface | Material | Cleaning schedule | Cleaning material | Who touches |
| --- | --- | --- | --- | --- |
| Inside floor | Vinyl tile | Variable, infrequent (~1/week) | Bleach | Universal – health care workers, visitors, patient if ambulatory |
| Outside floor | Vinyl tile | Variable, infrequent (~1/week) | Bleach | Universal – health care workers, visitors, patient if ambulatory |
| Inside door handle | Plastic in ICU; Steel outside ICU | Variable, infrequent (~1/week) | Hydrogen peroxide wipes | Universal – health care workers, visitors |
| Outside door handle | Plastic in ICU; Steel outside ICU | Variable, infrequent (~1/week) | Hydrogen peroxide wipes | Universal – health care workers, visitors |
| Bed Rail | Plastic | Variable, health care workers wipe down intermittently typically once at the start of shift (~2x daily) | Hydrogen peroxide wipes | health care workers, patient |
| Keyboard | Plastic | Variable, health care workers wipe down intermittently typically once at the start of shift (~2x daily) | Hydrogen peroxide wipes | health care workers |
| Air vent intake | Plastic | Variable, infrequent (~1/week) | Hydrogen peroxide wipes | health care workers |
| Ventilator buttons | Plastic | Variable, will be wiped down after no longer needed by patient (average 2-3 times a week) | Hydrogen peroxide wipes | health care workers (specifically respiratory therapists, MD) |
| Toilet seat | Ceramic | Variable, at least deep cleaned after patient discharged (average 2-3 times a week) | Bleach | Patient, visitors |

**Data file S1**. Statistical analysis of pairwise differences in log-ratio across sample types from figure 3D trajectory plot.

**Data file S2**. Top 100 random forest importance ranks and GreenGenes taxonomy from nares, forehead, stool, and inside floor samples.

## Notes

### Competing Interest Statement

The authors have declared no competing interest.

### Author Declarations

All participants were consented under UCSD Human Research Protections Program protocol 200613.

